# Determinants of Exerciser Environmentalism to Lower Meat Consumption in the Practice of Climate Stewardship

**DOI:** 10.1101/2025.10.19.25338270

**Authors:** Sean Kardar

## Abstract

**Introduction:** Animal husbandry for human consumption is a contributing factor of greenhouse gas accumulation. As country-sustainable dietary guidelines recommend, reducing meat overconsumption can substantially mitigate world greenhouse gas emissions. Globally, more than 35 years of evidence indicates a lack of awareness, willingness to alter dietary habits, and disbelief that meat consumption contributes to unwanted climate change.

**Methods:** Six hundred participants (exercisers, *n* = 300; nonexercisers, *n* = 300) were sampled randomly from the crowdsourcing database of Prolific and stratified across the age groups of 18–27, 28–37, 38–47, 48–57, and 58–100 with 30 males and 30 females per age group. Differences between exercisers and nonexcercisers were examined for environmental value orientations, beliefs, norms, and intentions related to climate degradation awareness and environmental friendliness in the context of an integrated value belief norm theory (VBN), theory of planned behavior (TPB), and health concern variable (VBNTPB) model.

**Results:** The results of three VBNTPB causal pathways indicated that exercisers differed significantly from nonexercisers, as compared to the VBN causal pathway, in predicting climate stewardship intention to reduce meat consumption when controlling for the sociodemographic factors of age, ethnicity, and education. Each VBNTPB causal pathway variable was positively and significantly related to the next, confirming an environmentalism pathway originating from biospheric, altruistic, and egoistic environmental value orientations. These values determined general proenvironmental worldview beliefs, which in turn predicted behavior-specific human meat reduction beliefs for awareness of consequences, ascription of responsibility, the moral obligation of personal norm, normative belief of social norm, and the behavioral beliefs of attitude and health concern, to conclude intention of lowering meat consumption to benefit Earth’s climate, and explained 85.4–87.3% of intention variance comparative to 83.2% in the VBN causal pathway. The additional variance explanation via attitude, health concern, and social norm in the VBNTPB model overcame the nonsignificant personal norm prediction of intention in the VBN causal pathway.

The common language effect size for each causal regression analysis indicated that exercisers and females would have a higher value on the outcome variable, ranging from 51.37%–72.87% and 50.92%–53.93%, respectively, when holding the values of other predictor variables constant, indicating a large effect size. Exercisers’ current behavior was 100% more likely to consume a low-meat diet than nonexercisers to benefit Earth’s climate (*OR* = 2.04).

**Conclusion:** The VBNTPB model is more effective in predicting climate stewardship. The outcomes indicate that individuals who regularly exercise within the range recommended by the U.S. Health and Human Services and the World Health Organization, to sustain the best health, are a worldwide population that society may leverage—to spur social movement in consuming less meat to prevent unwanted climate change. The rational evaluation of the behavioral beliefs of attitude and health concern, and the normative belief of social norm, are important behavioral antecedents that influence physically active individuals to overcome inadequate personal norm to conduct climate stewardship through dietary modification. These antecedents in the context of self-interest should be fostered and reinforced in humankind to promote environmentalism. Sociodemographic trends indicate that these antecedents should be taught throughout an individual’s interest in pursuing higher levels of educational credentials, at an early age, and throughout life’s progression.

## Introduction

Unnatural climate change is a historical and ongoing environmental problem. Earth’s temperature has increased by an average of 1.1 degrees Celsius from 1880 to the present (Lindsey & Dahlman, 2024; NASA, 2024), resulting in increased surface temperatures, rising sea levels, droughts, and other degrading ecosystem effects (WHO, 2023). The cause is predominantly due to anthropogenic sources that emit high amounts of carbon dioxide (CO_2_), nitrous oxide (N_2_O), and methane (CH_4_), collectively termed greenhouse gases (GHGs). Because human activity is the most significant source, it is the primary target for reducing greenhouse gas (GhG) emissions (Lindsey & Dahlman, 2024).

The future of humankind’s natural environment (Marinova & Bogueva, 2019) depends on mitigating ongoing temperature increases to maintain normative balances of human physiology. Increased illness and death are attributed to climate change-related heatwaves (Vicedo-Cabrera et al., 2021; Wallace et al., 2019; WHO, 2023). Vicedo-Cabrera et al. (2021) estimate that 37% of heat-related deaths across 732 countries stem from anthropogenic causes of climate change. Globally, human meat consumption has been increasing since the 1960s. If current rates continue, the demand for industrial meat and population growth will likely surpass ecosystem capacities, leading to a less habitable planet (Gonzắles et al., 2020; Hallstrom et al., 2015; Marinova & Bogueva, 2019; Meyer & Reguant-Closa, 2017).

Agricultural processes to rear livestock and related meat production generate 60-78% percent of GhGs produced by the food sector, accounting for 35-37% of all GhGs worldwide (EPA, 2025; Hallstrom et al., 2015; Meyer & Reguani-Closa, 2017; Mossburger et al., 2023; Xu et al., 2021). A simple act of meat reduction is a formidable resolution. However, this remains a complex social issue and a source of dissonance (Sanchez-Sabate et al., 2019; Sanchez-Sabate & Sabate, 2019). As the world population grows, GhG emissions from agricultural sources are likely to continue increasing.

Even with agricultural technological advances, only avoidance and reduced meat consumption are sufficiently effective to protect, sustain, and renew Earth’s deteriorating climate spurred by GhG’s (Gonzắles et al., 2020; Hallstrom, 2015; Macdiarmid, 2013; Myer & Reguant-Closa, 2017; Tilman & Clark, 2014; van Vliet et al., 2020). As reported in previous studies, the general public’s curbing of meat consumption is inadequate to comply with national and world dietary guidelines (Begue & Treich, 2019; Food and Agricultural Organization & World Health Organization [FAO & WHO], 2019; United States Department of Agriculture [USDA], 2020), and for environmental stewardship reasons (Sanchez-Sabate et al., 2019; Sanchez-Sabete & Sabete, 2019). Strategically, lowering the consumption of meat worldwide will substantially diminish GhG emissions. Vegetarian, Mediterranean, and Pescatarian alternative meat diets are estimated to reduce GhG emissions by as much as 55% (Gibbs & Cappuccio, 2021; Li et al., 2024; Tilman & Clark, 2014). Despite the high rate and declaration by 49% of individuals across 50 countries that climate change has reached an emergency status, and 70% of those individuals support exhausting any necessary corrective actions, support for a plant-based diet was the least popular (UNDP, 2021).

Worldwide studies across Europe and the United States continually find limited awareness of the causal connection between human meat consumption and unwanted climate change. Awareness levels range between 23–35%, and willingness to consume less meat after provisional education techniques range between 5–80% (Sanchez-Sabete & Sabete, 2019). The low proportion of awareness and large variability in willingness to consume less meat are attributed to limited knowledge of climate degradation outcomes, limited interest in carrying out climate stewardship, and disbelief that diet is causal to climate degradation. Individuals aware of the link between meat consumption and climate change tend to follow other climate stewardship practices. These practices include purchasing local produce, reducing the consumption of food items packaged in plastic, and advocating for the prevention of animal suffering (Lea & Worsley, 2008; Vanhonacker et al., 2013). The motives of participants prepared for dietary change aligned better with health maintenance and the naturalness of food consumed (Lea & Worsley, 2008; Macdiarmid et al., 2016; Macdiarmid, 2013).

Intervention efforts to raise awareness and willingness have primarily targeted information provisioning, message framing, and cognitive dissonance (Graham & Abrahamse, 2017; Hunter & Röös, 2016; Kunst & Haugestad, 2018; Kunst & Hohle, 2016; Malek et al., 2019; Pabian et al., 2020; Kwasny et al., 2021). Despite the moderately successful outcomes, the continuous and widespread facilitation of these methods is impractical. Individuals must consent to receiving intervention content and commit to regular viewing. Since the world’s population is generally unwilling to limit meat consumption for environmental benefit, accepting psychologically-based content to alter their mindset is unlikely.

Ensuring population awareness of the causal link and outcome is requisite to minimize the problem–awareness and value-action gaps that prohibit choosing low-meat diets to prevent climate degradation caused by meat overconsumption (Kwasny et al., 2021; Weibel et al., 2019). A less direct approach may better serve individuals unaware or aware but remain unwilling to alter their diet. The behavioral influences of why humans resist consuming a low-meat diet to benefit Earth’s climate were the focus of this study. The current literature primarily focuses on external interventions (Bianchi et al., 2018; Ronto et al., 2022). Behavioral change models are useful for understanding the antecedents to dietary change. These models depict the antecedents of human behavior against transitional steps leading to behavioral change (Borusiak et al.,2021; Carfora et al., 2019; Lai et al., 2020). Evaluating inherent behavioral antecedents that support climate stewardship to follow a low-meat diet is warranted to promote voluntary dietary change.

Because no single theory includes all the constructs known to be operation to facilitate behavioral change needed to affect climate degradation, an integrated value-belief-norm theory (VBN), theory of planned behavior (TPB), and health concern variable theoretical model is needed, termed here as the VBNTPB model. Each theory and health concern variable is relevant to develop the necessary constructs to examine intervention strategies for humans to consume a low-meat diet.

Biospheric, altruistic, and egoistic value orientation constructs are known antecedents to environmentally friendly behavior (Ahmat et al., 2022; deGroot & Steg, 2010; Marshall et al., 2019). These values mediated environmental concern, indicating that values influence an individual’s beliefs related to awareness of the consequences and their ascription of responsibility to offset climate degradation (Hansla et al., 2008). As VBN values develop through life experience, an individual’s awareness of consequences aligns with their VBN proenvironmental values, enabling them to initiate action to resolve adverse environmental impacts. Their efforts to address the negative consequences stem from their sense of responsibility.

In support, Kovacevia et al. (2024) found that individual decisions to hold others responsible for their actions depend on whether the person facilitating the action should or could have easily known the outcomes were deleterious and should have made alternative decisions. Kaiser and Shimoda (1999) explained how feelings of guilt account for a large proportion of whether individuals feel responsible for their actions. The collective findings indicate that awareness and understanding are necessary for individuals to accept responsibility.

Attitude is a prerequisite to proenvironmental behavior. Miller et al. (2022) found that an individual’s attitude across 11 countries was positively associated with proenvironmental behavior. Additionally, an environmental attitude facilitates the acquisition of information, making individuals more environmentally aware and more likely to exhibit proenvironmental behavior as ecological conditions worsen (Baleri et al., 2022; Wyss et al., 2022). Attitudes are ending evaluations of topics and ideas and describe personal feelings derived from life perspectives (Borusiak et al., 2021; Darnton, 2008; Kwasny et al., 2021). They are a frame of reference summed by criteria such as beliefs, social interaction, rearing, and education (Ajzen, 2020; Brusiak et al., 2021; Darnton, 2008).

A proenvironmental worldview is important to help operationalize a proclimate attitude and behavior. The broad proenvironmental worldview beliefs of human actions on the environment influence individuals’ acceptance of more specific awareness of consequences and outcomes. Ballew et al. (2019) found that when individuals believe humanity and nature are connected as one and interdependent, it explains their beliefs and attitudes about preventing unwanted climate change. Prenvironmental worldview beliefs explain proenvironmental behavior (Derdowski, 2020).

An individual’s concern for health is a likely reason to consume less meat and concurrently benefit Earth’s climate (Battaglia et al., 2015). Yet, Tallie et al. (2022) reported that environmental concern messages to preserve the environment were more effective than health only messages to reduce meat consumption. Comparatively, Neff et al. (2018) found the opposite finding. Individuals were willing to reduce meat consumption for health benefits but less to remedy environmental concerns. A concurrent evaluation of protecting the environment and concern for health is necessary to understand if health concerns influence dietary change to a low-meat diet in the context of climate stewardship.

Proenvironmental behavior is also an outcome of a moral responsibility that is a product of personal norms. Personal norms correlate with proenvironmental behavior across multiple studies (Bai et al., 2020; Niu et al., 2023; Pearce et al., 2022; Tran et al., 2020). Braksiek et al. (2021) identified significant correlations among the theory of planned behavior (TPB) antecedents of attitude, perceived behavior control, social norms, and intention on proenvironmental behavior for sports club members. Hence, an individual’s settled way of thinking, perceived behavioral control to overcome barriers, and the influence of others who are considered important influence physically active individuals’ development of proenvironmental behavior.

Collectively, the literature findings suggest that if an individual has an inherent point of view based on their biospheric, altruistic, and egoistic value orientations, proenvironmental worldview, awareness of consequences, ascription of responsibility, attitude, perceived behavioral control, personal and social norms, health concerns, and intention, they will be more likely to practice climate stewardship. Thus, by evaluating the influences of each construct in determining proenvironmentalism, a model including all these constructs is necessary to determine how they influence an individuals’ decision to consume less meat to reduce climate degradation. Combining the constructs of the VBN, TPB theories and a health concern variable will allow a full investigation of the determinants of dietary behavior change.

Minimizing climate degradation by consuming less meat is a personal commitment. Its success could reduce meat-related GhG emissions between 30–55% depending on societal compliance (Gibbs & Cappuccio, 2020; Rust et al., 2020; Tilman & Clark, 2014). Individuals fail to develop corrective intentions or behavior even after acknowledging the connection. A lack of concern and disinterest indicates motivation is lacking to encourage voluntary diet change, indicating moral disengagement exists prohibiting climate stewardship actions (Heald, 2017; Sanchez-Sabete et al., 2019).

Physically active individuals believe climate degradation affects them personally, support better climate policy, and demonstrate inherent proenvironmental behavior (Cunningham et al., 2020; Braksiek et al., 2021; Thorman & Wicker, 2021). These physically active individuals may also exhibit traits of altruism and egotism (Ortega-Egea et al., 2014). Other studies show the existence or development of similar characteristics in physically active people (Buragohain & Senapati, 2016; Duygu et al., 2019; Kim & Ahn, 2021). Following the VBN theory, those who meet a certain level of planned physical activity, or “exercisers,” will feel obligated to protect what is considered valuable (Stern et al., 1999). For these reasons, exercisers are more likely to lower their meat consumption.

If people who exercise possess increased proenvironmental views, this group may be more likely to reduce their meat consumption and encourage others similarly. The outcomes could elevate social practices to improve the climate, termed climate stewardship, specifically, a change in diet to lower meat consumption. Exercisers are a likely group of individuals to lead behavior change related to lower meat consumption because of inherent values to practice climate stewardship. The most relevant constructs of behavior change manifest in this group compared to individuals who do not exercise regularly are unknown. Therefore, the exact points of intervention in the proposed VBNTPB model of behavior change cannot be targeted for optimal intervention.

A study was undertaken in a group of exercisers and nonexercisers to investigate 12 constructs representing latent concepts in the VBNTPB model to determine which are ideal for designing successful interventions to lower meat consumption for reducing climate degradation. Additionally, differences between exercisers and nonexercisers willingness and behavior to modify diet by reducing meat consumption were evaluated. This study provides a foundation for possible new approaches to increase awareness of the connection between meat consumption and climate degradation.

The following subscale and objective research hypotheses were tested:

**H1**: Exercisers biospheric value orientation positively influences proenvironmental worldview more than nonexercisers.
**H2**: Exercisers altruistic value orientation positively influences proenvironmental worldview more than nonexercisers.
**H3**: Exercisers egoistic value orientation orientation positively influences proenvironmental worldview more than nonexercisers. .
**H4**: Exercisers proenvironmental worldview positively influences awareness of consequences more than nonexercisers.
**H5:** Exercisers awareness of the consequences of unnatural climate change caused by human meat consumption positively influences ascription of responsibility more than nonexercisers.
**H6:** Exercisers ascription of responsibility to protect Earth’s climate toward consuming less meat positively influences personal norm more than nonexercisers.
**H7:** Exercisers personal norm to protect Earth’s climate toward consuming less meat positively influences intention more than nonexercisers.
**H8**: Exercisers have a more climate friendly attitude toward reducing meat consumption to protect Earth’s climate positively influences intention more than nonexercisers.
**H9:** Exercisers perceived behavioral control to protect Earth’s climate toward consuming less meat positively influences intention more than nonexercisers.
**H10:** Exercisers social norm to protect Earth’s climate toward consuming less meat positively influences intention more than nonexercisers.
**H11:** Exercisers health concern to protect Earth’s climate toward consuming less meat positively influences intention more than nonexercisers.
**H12:** Exercises frequency in following a modified diet to lessen unnatural climate change will exceed nonexercisers.

## Methods

A.T. Still University Institutional Review Board approved this study, protocol # SK20230323-001 (S1 File). A causal-comparative research design was used to survey a U.S. sample classified as exercisers and nonexercisers. Individuals qualifying as exercisers attested to completing a minimum of 60 minutes of physical exercise weekly (Prolific, 2023; U.S. Department Of Health & Human Services, 2024). This requirement is in the range of the U.S. Health and Human Services weekly physical activity guideline recommendation of 75 or 150 minutes of vigorous or moderate exercise. The comparative nonexerciser group was anyone who attested to less than 60 minutes of exercise per week.

### Survey Formatting

A novel survey was created by the researcher based on an integrated VBNTPB model (S2 File). The survey contained 63 items comprising 12 construct groups and one objective current behavior item. Three items were screening questions, and 60 were reflective, Likert items. Construct item groups from the environmental portrait value questionnaire (EPVQ) and the new ecological paradigm (NEP) scale measured values and proenviornmental worldview and accounted for 14 and six survey reflective items, respectively (Anderson, 2012; Dunlap, 2000; Bouman et al., 2018). The EPVQ scale contained four reflective items to measure participants’ inherent biospheric value orientation, five to measure altruistic value orientation, and five to measure egoistic value orientation. The NEP scale included six items to measure participants’ proenvironmental worldview.

The remaining reflective items measured participants’ responses for seven constructs: awareness of consequences, ascription of responsibility, attitude, personal norm, perceived behavioral control, social norm, and health concern, containing five items each—one neutral, three positive, and one negative. Two additional items were added to the survey to measure the intention of participants to change their diet, and their current meat consumption related to climate improvement.

The structure of the researcher-created reflective survey items followed the current literature and direction described by Ajzen (Ajzen, n.d.; Ajzen, 2006). The following surveys measured previously for reliability and validity were used as a guide to create the survey items: Alam et al. (2020); Borusiak et al. (2021); Carfora et al., 2019; Coker & van der Linden, 2020; Gilford et al., 2022; Hoeksma et al. (2017); Lai et al. (2020), and Tommasetti et al. (2018). The psychometric quality and survey item content validity is subjective and judgmental. Extensive research was completed to inspect other related survey items, formats, and content for the VBN and TPB theories, and health concern predictor. The researcher-developed survey items were developed iteratively with ATSU advisers. Feedback was also solicited from other ATSU personnel (faculty and staff) and other pilot survey participants.

### Sample Description

Study participants were recruited through the world’s largest crowdsourcing research platform, Prolific. All participants provided online written informed consent. Prolific sent emails to selected members based on parameters provided by the researcher. Participants were required to be fluent in English and currently living in the United States (verified via I.P. address). Strata were constructed to provide equal numbers of exercisers and nonexercisers by sex (male or female), ethnicity (White, Mixed, Asian, Black, and Other), and age in years (18–27, 28–37, 38– 47, 48–57, and 58–100). Once the sample was identified, Prolific sent the survey to each participant via an emailed SurveyMonkey link.

Study sampling occurred between April 15–May 1, 2023. Over 1,000 Prolific members were invited to complete the survey. Of these, 488 exercisers and 536 nonexercisers entered the survey. Given that some participants entering the survey were disqualified from completing the survey, omitted or rejected from sample populations, the following attrition rates were identified. Participant survey completion times less than three standard deviations (6.90 minutes) of the average completion time (7.35 minutes) were rejected using Prolific’s rejection feature, resulting in the removal of surveys from 107 exercisers (21.92%) and 80 nonexercisers (14.92%). The average completion time used to reject participants was calculated from the 1st-day cohort of participants completing the causal comparative study. For both groups, the ending survey average completion time was 7.42 minutes on May 1, 2023. The average disqualifying nonconsenting rate of the consent form was .19% (S3 and S4 File).

### Data Cleaning

Exercisers and nonexercisers whose exercise habits did not match their Prolific profile as determined by a qualifying question after entering the survey were disqualified (7.39% and 25.25%, respectively). Exercisers (7.76%) whose physical activity matched their Prolific profile but indicated physical activity was not undertaken to maintain a good health status were omitted from the exercise sample population after completing the survey. The final completion rates were 20% for exercisers and 27% for nonexercisers.

The results of the data cleaning process ended with 300 exercisers and 300 nonexerciser participants who completed the study survey in 6.90 minutes or greater. All participants who completed the survey and were not disqualified or rejected were paid a nominal fee of $3 to complete the survey. Each group contained 30 males and 30 females per the age groups of 18–27, 28–37, 38–47, and 58–100. Construct scores for each participant were created by summing the Likert item responses assigned to each construct to create continuous variables for each construct for each participant.

### Latent Construct Variables

Based on the most favorable Cronbach’s alpha reliability of .7, the following survey items were summed to create continuous variables for each construct (S5 File): 1–4 (Biospheric), 5–9 (Altruistic), 10–12 (Egoistic), 15–20 (Proenvironmental view), 21–25 (Awareness of consequences), 26–30 (Ascription of responsibility), 31–35 (Attitude), 36–40 (Personal norm), 43–48 (Perceived behavioral control), 49––52 (Social norms), 54–57 (Health concern), and 58– 62 (Intention). The survey settings and definitions of the variables are located in the survey codebook (S6 File).

### Current Behavior Survey Question

For each exerciser and nonexerciser group, Questions 1 and 3 of the current dietary behavior items were combined to calculate the total number of participants who changed their diet, and Questions 4 and 5 were combined to calculate the total number of participants who did not change their diet (S7 and S8 File).

### Research Design

Research hypotheses for the 12 latent constructs were analyzed as a causal pathway reflective of the VBN and TPB theories with a health concern variable. Exercisers were postulated to have higher beta coefficients for the dependent variables than nonexercisers. Five sequential multivariate linear regression models were performed to predict dependent variable outcomes and adjust for differences between the exerciser and nonexerciser groups in terms of previous independent variables within the causal pathway, as well as age, sex, ethnicity, and education, when predicting the 12 subscales of the survey instrument and the total score.

Dummy-coded variables were created for the categorical control variables. The statistical assumptions of normality, linearity, homoscedasticity, autocorrelation, and multicollinearity were assessed before model interpretation. The unstandardized beta coefficients, their respective standard errors, and the standardized beta coefficients for the exerciser versus nonexerciser parameter related to each block of the sequential regression analyses were reported.

The exerciser versus nonexerciser variable (nonexerciser was used as the reference) was entered into the models along with age (18-27 = reference, 28-37 = 1, 38-47 = 2, 48-57 = 3, and 58-100 = 4), sex (male = reference, female = 1), ethnicity (White = 0, Asian = 1, Black = 2, Mixed = 3, and Other = 4), and education (undergraduate/graduate school = reference, technical/community college = 1, secondary = 2, no formal education = 3, and unknown = 4) when predicting for the subscale and total outcomes associated with the survey instrument. The statistical assumptions of each model were assessed and met.

Each regression analysis variable was measured within the following acceptable ranges (DW = 1.5-2.5, T = >.1 and < 1.0, VIF = < 10). Statistical significance was assumed at a one and two-sided alpha values of .05 or less, and all analyses were performed using SPSS Version 29 and 30 (IBM Corp.2024).

## Results

Using Prolific’s simplified ethnicity categorization, the majority of the study participants, exerciser and nonexerciser, respectively, were White, resided in the U.S. South regional location, and held an undergraduate degree as their highest level of education. The exerciser group comprised 19.67% fewer high school graduates as their highest level of education but 11.67% and 5.34% more undergraduate and graduate credentialed participants (Table 1). Table 1 shows the exercise group consisted of 141 (47%) who exercised sometimes (60–150 minutes per week) and 159 (53%) who exercised often (> 150 minutes per week).

**Table 1.** Participant Demographic Data.

| Criteria | <u>Exerciser Study Group (n = 300)</u> | <u>Nonexerciser Study Group (n = 300)</u> |
| --- | --- | --- |
|  | Percent of Total (%) | Percent of Total (%) |
| Ethnicity |  |  |
| White | 81.33 | 75 |
| Black | 5.33 | 11.33 |
| Asian | 3.66 | 4.33 |
| Other | 4.33 | 4.00 |
| U.S. Region |  |  |
| South | 40.66 | 43.33 |
| Midwest | 21.33 | 21.00 |
| Northeast | 24.66 | 19.33 |
| West | 13.33 | 16.33 |
| Education |  |  |
| Graduate degree<br>(M.A./M.Sc/MPhil/Other) | 13.00 | 7.66 |
| Undergraduate<br>(B.A./BSc/Other) | 37.33 | 25.66 |
| Technical & community<br>college | 20.66 | 19.66 |
| High school diploma | 21.66 | 41.00 |
| Secondary education<br>(GED/GSCE) | 2.33 | 4.00 |
| No formal education | 1.00 | 0.66 |
| Frequency of Exercise |  |  |
| Often (> 150 minutes<br>weekly) | 47.00 |  |
| Sometimes (60—150<br>minutes weekly) | 53.00 |  |
| Never (0–60 minutes<br>weekly) |  | 100 |

The following univariate statistical analyses described the exerciser and nonexerciser data: frequency, standard deviation, interquartile range, and median values of the continuous variable Likert scores for each exerciser and nonexerciser group. The mean, standard deviation, median, and interquartile range were used to describe the derived constructs (S9 File). Median constructs generally ranged between 20–31 (*IQR* = 4–11). The egoistic and social norm constructs were somewhat lower than other constructs for both exercisers, 8(*IQR* = 5) and 14.5(*IQR* = 6), and nonexercisers, 8(*IQR* = 5) and 13(*IQR* = 6). The values indicate that participants were less self-centered and experienced less influence from their environment to consume a low-meat diet to reduce unwanted climate change.

The standard deviations of the constructs ranged between 0.01–0.44, indicating minimal variance, and Tukey Fence outliers ranged between 0–3.34% for each construct (S9 and S10 Files). Exercisers’ and nonexercisers’ construct scores did not differ more than 2.2%. Each construct contained outliers except attitude, personal norms, perceived behavioral control, and intention. The minimal outliers remained in the dataset as natural variations of data collection.

### Validity and Reliability

The meaningfulness of the causal comparative study relies on adequate reliability and validity data from both exerciser and nonexerciser study populations. An assessment of the data construct validity via confirmatory factor analysis (CFA) does not reflect a matrix showing survey subscale items to load separately on each of the survey scale’s 12 constructs due to multiple cross loading of items, a known limitation of closely related items (Muller-Schneider, 2022; Watkins, 2018).

The construct validity of the researcher-created survey is confirmable by known-groups validity—when a test or questionnaire can discriminate between groups known to differ on a variable of interest and grounded in robust and evidentiary findings. In this study, the known groups are individuals who complete more physical activity than others who do not, and the variable of interest is environmentalism.

The literature supports defining more physically active individuals and their inclination to carry out proenvironmental behavior as a known group. In support, Cunningham et al. (2020) found that as physical activity increased, so did people’s belief that climate change personally affected them. They also concurrently found increased support for new climate policies compared to sedentary and less physically active individuals. Teixeria et al. (2023) reported that physical activity is associated with proenvironmental behavior, and increased physical activity found a more significant association.

Thorman and Wicker (2022) found that sports club members were more willing to pay a carbon tax to offset global warming, which increased positively with their environmental consciousness scores and weekly sports practice hours. In a subsequent study, Thorman and Wicker (2022) found that sports club members’ environmental consciousness score is positively associated with proenvironmental actions and negatively associated with their carbon footprint.

This study further defined physical activity as those who complete exercise to maintain good health. The qualification minimizes the ambiguity of physical activity, limiting it exclusively to exercise to benefit health. The developed survey examined behavioral antecedents, willingness, and current behavior to modify diet by consuming less meat to benefit Earth’s climate comparable to nonexercisers.

The survey outcome of Figure 1 justified the known groups’ qualification by discriminating between exercisers’ and nonexercisers’ environmentalism. The aggregate score across the survey behavioral predictors between the groups was statistically significant when predicting the final causal pathway outcome variable of intention, *p* = 0.01, and a significant chi-square test of association for diet modification was also concluded, *p* < 0.001 (Figure 3). The significant intention prediction indicates predictive validity of the survey underlying dimensionality of environmentalism willingness (VIF = 1.0-1.93, T = .58-.97, DW = 2.06).

**Figure 1.**
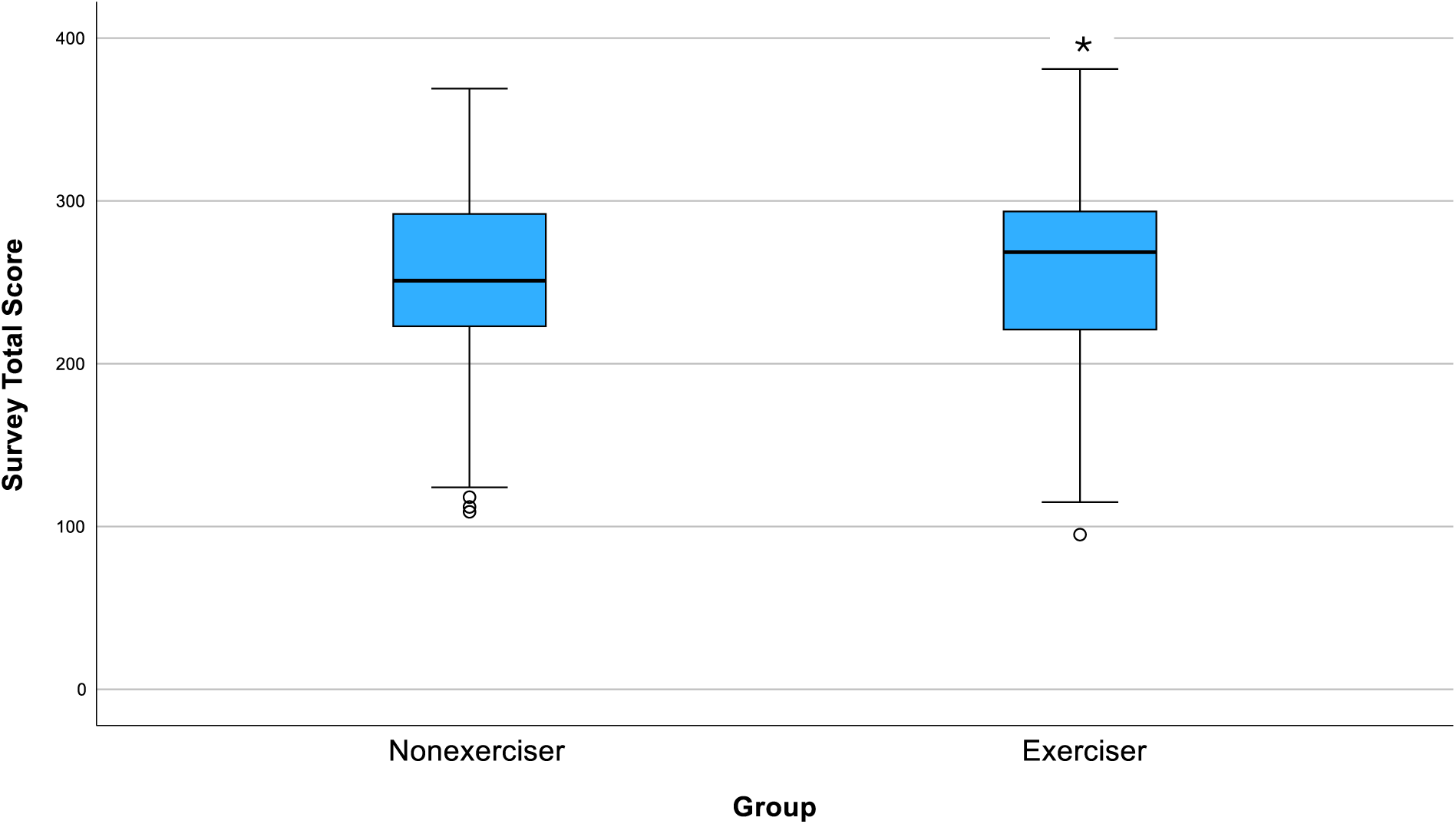
Difference in Survey Total Score when Predicting Intention.

Reliability of the survey was assessed with the collected data. The data indicate that each survey construct is reliable in reproducing the same outcomes externally beyond the study, for which the measurements were values, beliefs, norms, concerns, and intentions. Individual construct reliability ranged from moderate (α = .67) for social norms to excellent (α = .95) for awareness of consequences.

### Statistical Conclusion Validity

The data exhibited exceptional statistical conclusion validity defined by a comprehensive measure of study power, significance testing, and effect size. Across all constructs, the survey power is 99.9%, and the exerciser group common language effect sizes ranged between 51.37%–72.87% (Faul et al., 2007; Hernandez, n.d.; Krasikova et al., 2018; Vargha & Delaney, 2000).

### Research Findings

A series of sequential regression analyses were conducted based on an integrated value belief norm theory (VBN), theory of planned behavior (TPB), and health concern variable (VBNTPB) model.

**Figure 2.**
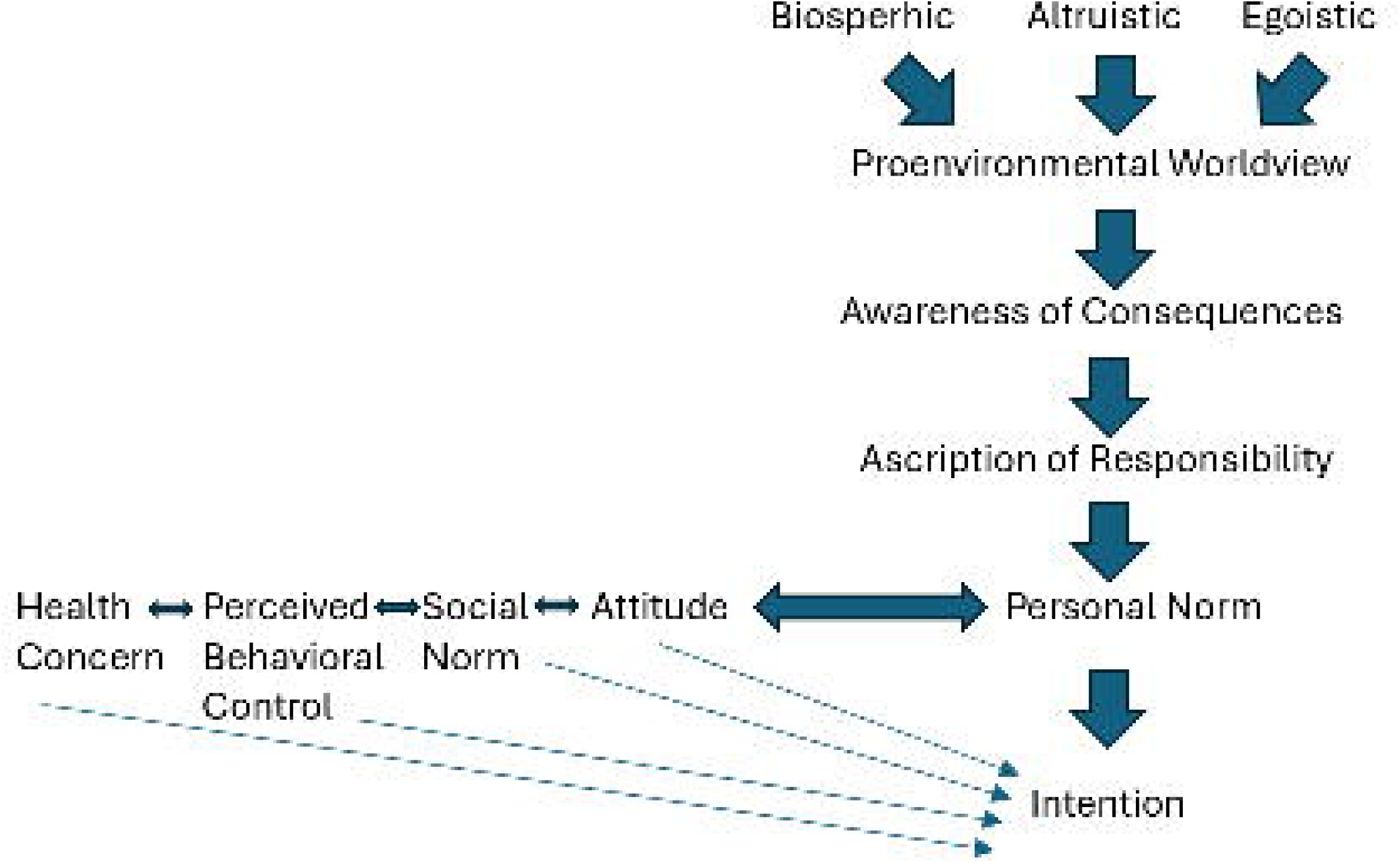
VBNTPB Causal Pathway Model.

Each dependent variable of the causal pathway was regressed onto the preceding predictor variables in the pathway. After each model regression analysis, the dependent variable was added to the existing predictor variables, and the next outcome variable of the causal pathway was selected as the dependent variable. This iterative process concluded with intention as the final outcome variable. Predictor variables for the theory of planned behavior (TPB) and health concern were added at the level of personal norm to form an integrated VBNTPB model. The Bonferroni calculation was applied to offset the increased likelihood of a Type I error when conducting multiple analyses. The adjusted significance threshold for the models was set at *p* < 0.01, calculated by .05 divided by 5 regression analyses. (Hoeksma et al., 2017; Sahin, E., 2013; Steg et al., 2005). Each regression model resulted in a significance level with a *p*-value of less than 0.001.

Hypotheses 1-7 were confirmed for the VBN causal pathway, and Hypotheses 1-7 and 8-11 for the VBNTPB across three distinct causal pathways. However, hypothesis 9 was not statistically significant for exercisers, and the data are not presented. Hypothesis 12 was confirmed.

### VBN Causal Pathway (S11_Table 2)

Value orientation variables were the predicting variables in block one, and proenvironmental worldview was the dependent variable. The adjusted R-square of the model indicated that value orientations were statistically significant and explained 30.2% of proenvironmental worldview variance (*p* < .001). Exercisers’ (b = 1.22, β = .07, *p* = .04, *partial correlation* = .08) and females’ (b = 1.43, β = .08, *p* = .01, *partial correlation* = .09) biospheric (β = .40, *partial correlation* = .36, *p* <.001), altruistic (β = .15, *partial correlation* = .15, *p* <.001), and egoistic (β = .07, *partial correlation* = .08, *p* = .04) value orientations were positive and significantly associated with proenvironmental worldview when controlling for all predictor variables, the biospheric value orientation was the most significant unique contributor (*partial correlation* = .36).

A one-unit increase in exercise status and gender was associated with a 122% and 143% increase in proenvironmental worldview for exercisers and females. Indicating the stronger exercisers’ and females’ biospheric, altruistic, and egoistic value orientations, the stronger their belief that natural resources should be protected and misuse by humans can negatively impact nature. The common language effect size (CLES) indicates that a randomly selected exerciser and female with higher biospheric, altruistic, and egoistic value orientations will also have a higher proenvironmental worldview, 52.65% and 53.16% more than nonexercisers or males, when values on other predictors are the same.

All age groups were positively and nonsignificantly associated with a proenvironmental worldview. Aside from the Mixed ethnic category (β = -.04, *partial correlation* = -.05, *p* = .22), which was negatively associated with proenvironmental worldview, all other ethnicities were positively associated, and Asians (β = .07, *partial correlation* = .08, *p* = .03) were statistically significantly different. Every level of education was negatively associated with participants’ proenvironmental worldview compared to those achieving higher education; only secondary level education (β = -.07, *partial correlation* = -.08, *p* = .04) was statistically significant.

In block two, awareness of consequences was the dependent variable, and the predictor variables were proenvironmental worldview and the value orientations of altruistic, egoistic, and biospheric. The adjusted R-square of the model indicates that the predictor variables explained 42.2% of the awareness of consequences variance, was significant (*p* <.001), and increased the VBN causal pathway explanation 11.8%. Proenvironmental worldview (β = .46, *partial correlation* = .41, *p* <.001) was the most unique and significant contributor to predicting awareness of consequences.

Exercisers’ (b = 1.73, β = .10, *partial correlation =* .12, *p* = .00) and females’ (b = .661, β = .03, *partial correlation =* .05, *p* = .23) proenvironmental worldview positively associated with awareness of consequences. Exercisers were statistically significantly different than nonexercisers. A one-unit increase in exercise status and gender was associated with a 173% and 66.1% increase in proenvironmental worldview for exercisers and females. Therefore, exercisers and females demonstrated stronger awareness of the consequences for harmful outcomes to Earth’s climate caused by greenhouse gas due to human meat consumption as their proenvironmental worldview increased, compared to nonexercisers and males. The CLES indicates that a randomly selected exerciser and female with a higher proenvironmental worldview will also have a higher awareness of consequences, 54.05% and 51.59% more than nonexercisers or males, when values on other predictors are the same.

The value orientations of biospheric (β = .18, *partial correlation* = .17. *p* <.001) and egoistic (β = .11, *partial correlation* = .14, *p* <.001) were positive and statistically significant. The altruistic value orientation (β = .06, *partial correlation* = .06, *p* = .12) was positively associated and nonsignificant.

The influence of age on awareness of consequences was statistically nonsignificant and increased positively from younger to older; age groups 48-57 (β = .01 *partial correlation* = .01, *p* = .71) and 58-100 (β = .04, *partial correlation* = .03, *p* = .34) were the most impactful. The Mixed (β = -.05, *partial correlation* = -.06, *p* = .09) ethnicity was negatively and nonsignificantly associated with awareness of consequences, and the remaining ethnic groups were positive and statistically nonsignificant. Across all levels of education, participant responses were negatively associated with awareness of consequences, and only secondary education was statistically significant (β = -.08, *partial correlation* = -.09, *p* = .02).

In the third block, the ascription of responsibility was the dependent variable, and the predictor variables were awareness of consequences, proenvironmental worldview, and the value orientations of altruistic, egoistic, and biospheric. The adjusted R-square of the model indicated 57.6% of the ascription of responsibility variance was explained, significant (*p* <.001), and increased the VBN causal pathway explanation 15%. Awareness of consequences (β = .59, *partial correlation* = .51, *p* <.001) was the most unique and significant contributor to predicting ascription of responsibility.

Exercisers’ (b = 1.24, β = .07*, partial correlation =* .10, *p* = .01) and females’ (b = 1.42, β = .08, *partial correlation =* .12, *p* = .00) ascription of responsibility positively and statistically significantly increased as their awareness of consequences increased. A one-unit increase in exercise status and gender was associated with a 124% and 142% increase in awareness of consequences for exercisers and females. As exercisers’ and females’ awareness of consequences increased, the stronger their ascription of responsibility belief that human actions to lessen the consumption of meat can avert harm to Earth’s climate due to greenhouse gas compared to nonexercisers and males. The CLES indicates that a randomly selected exerciser and female with a higher awareness of consequences will also have a higher ascription of responsibility, 53.41% and 53.93% more than nonexercisers or males, when values on other predictors are the same.

Participants’ biospheric (β = .15, *partial correlation* = .16, *p* < .001) and egoistic (β = .11, partial correlation = .17, p < .001) value orientations positively and significantly predicted their ascription of responsibility. Constratingly, proenvironmental worldview was positive (β = .08, *partial correlation* = .07, *p* = .06), and the altruistic value orientation (β = -.01, *partial correlation* = -.01, *p* = .72) was negative; both were nonsignificant.

The influence of age on awareness of consequences was positively related and increased from younger to older. The 48-57 (β = .08, *partial correlation* = .09, *p* = .02), and 58-100 (β = .10, *partial correlation* = .11, *p* = .00), age groups were statistically significant. Blacks (β = .06, *partial correlation* = .08, *p* = .03) were positively and significantly associated with an awareness of responsibility; the remaining ethnicities were only positively associated, except the Mixed (β = -.03, *partial correlation* = -.04, *p* = .27) participants, who were negatively associated. All levels of education were statistically nonsignificant and primarily negatively associated with an ascription of responsibility. Technical/Community college was positively associated (β = .01, *partial correlation* = .02, *p* = .56).

In the fourth block of the VBN causal pathway, personal norms were the dependent variable, and the predictor variables were the ascription of responsibility, awareness of consequences, proenvironmental worldview, and the value orientations of altruistic, egoistic, and biospheric. The adjusted R-square indicated that the model explained 65.1% of personal norm variance, was statistically significant (*p* <.001), and increased the VBN casual pathway explanation 7.3%. Ascription of responsibility (β = .44, *partial correlation* = .42, *p* <.001) was the most unique and significant contributor to predicting personal norms.

Exercisers’ (b = 1.06, β = .06, *partial correlation =* .10, *p* = .01) and females’ (b = 1.14, β = .06, *partial correlation = .*11, *p* = .00) ascription of responsibility positively and significantly associated with personal norms. A one-unit increase in exercise status and gender was associated with a 106% and 114% increase in personal norm for exercisers and females. Thus, as exercisers’ and females’ ascription of responsibility increased, the stronger their personal norm to morally take action to lessen meat consumption to reduce harm to Earth’s climate due to greenhouse gas, compared to nonexercisers and males. The CLES indicates that a randomly selected exerciser and female with a higher ascription of responsibility will also have a higher personal norm, 53.22% and 53.51% more than nonexercisers or males, when values on other predictors are the same.

Participants’ biospheric (β = .14, *partial correlation* = .17, *p* <.001) and egoistic value orientations (β = .07, *partial correlation* = .12, *p* = .00) and awareness of consequences (β = .30, *partial correlation* = .27, *p* <.001) positively and significantly predicted personal norm when controlling for all other variables. In contrast, proenvironmental worldview (β = .02, *partial correlation* = .02, *p* = .51) was positive, and the altruistic value orientation (β = -.04, *partial correlation* = -.05, *p* = .20) was negative, both were nonsignificant.

The influence of age on awareness of consequences was statistically nonsignificant and increased negatively to positively concurrent with increasing age. The ethnic categories of Black (β = .04, *partial correlation* = .06, *p* = .10) and Other (β = .02, *partial correlation* = 04, *p* = .32) were positive, Asian (β = -.00, *partial correlation* = -.01, *p* = .76), and Mixed (β = -.02, *partial correlation* = -.04, *p* = .35) were negative; all groups were statiscally nonsignificant. Secondary level (β = .00, *partial correlation* = .01, *p* = .80) education participants were positively related, and the remaining education levels were negative; all were statistically nonsignificant.

In the final block of the VBN causal pathway, intention was the dependent variable, and personal norm, ascription of responsibility, awareness of consequences, proenvironmental worldview, and the biospheric, altruistic, and egoistic value orientations were the predictor variables. The adjusted R-square of the model indicated that 83.2% of the intention variance was significantly explained (p < .001), increasing the VBN causal pathway explanation 17.5%. Personal norm (β = .69, *partial correlation* = .72, *p* <.001) was the most unique and significant contributor to predicting intention.

Exercisers’ (b = .318, β = .01, *partial correlation* = .04, *p* = .29) and females’ (b = .457, β = .02, *partial correlation* = .06, *p* = .13) personal norm positively increased as their intention increased but was statistically nonsignificant. A one-unit increase in exercise status and gender was associated with a 31.8% and 45.7% increase in intention for exercisers and females. This means that as exercisers’ and females’ personal norm increased, their intention to lessen meat consumption to reduce harm to Earth’s climate due to greenhouse gases became stronger than nonexercisers and males. The CLES indicates that a randomly selected exerciser and female with a higher personal norm will also have a higher intention, 51.37% and 52.01% more than nonexercisers or males, when values on other predictors are the same.

Participants’ awareness of consequences (β = .11, *partial correlation* = .13, *p* <.001) and ascription of responsibility (β = .15, *partial correlation* = .20, *p* <.001) were positive and significantly associated with intention. Biospheric (β = .02, *partial correlation* = .04, *p* = .24) and egoistic value orientations (β = .01, *partial correlation* = .03, *p* = .42), and proenvironmental worldview (β = .01, *partial correlation* = .01, *p* = .67) were positive and nonsignificant. In contrast, the altruistic value orientation (β = -.03, *partial correlation* = -.06, *p* = .11) was negatively associated with intention and statistically nonsignificant.

All age groups were negatively and nonsignificantly associated with intention. The Mixed (β = -.01, *partial correlation* = -.04, *p* = .33) ethnicity was negatively related, and those remaining were positively related; all ethnicities were statistically nonsignificant. Each education level was negatively associated and primarily statistically nonsignificant; the level of no formal education (β = -.04, partial correlation = -.10, *p* = .00) was solely significant.

### VBNTPB Causal Pathway 1 (Attitude and Personal Norm) (S12_Table 3)

A significant effect in the VBNTPB causal pathway was identified when adjoining attitude with personal norm as predictors in block five. The VBNTPB casual pathway one differs from the VBN casual pathway in that intention was regressed onto attitude and personal norm, and increased the VBNTPB casual pathway one explanatory power 19.7%. The adjusted R-square of the model indicates the model predictors explained 85.4% of intention variance and was statistically significant (*p* <.001). Personal norm were the most impactful unique significant contributor (β = .44, *partial correlation* = .45, *p* <.001) to predicting intention.

Exercisers’ (b = .533, *partial correlation* = .07 *p* = .03, one-sided) and females’ (b = .391, *partial correlation =* .05, *p* = .16) attitude and personal norm were more positively associated with intention – exercisers were statistically significant, and females were not. A one-unit increase in exercise status and gender was associated with a 53.3% and 39.1% increase in intention for exercisers and females. The stronger the exercisers’ and females’ personal norm and attitude, the stronger their intention, an evaluation of whether participants intended to take action to lower meat consumption to help protect Earth’s climate caused by greenhouse gases. The CLES indicates that a randomly selected exerciser and female with higher attitude and personal norm will also have a higher intention, 52.49% and 51.85% more than nonexercisers or males.

Ascription of responsibility (β = .86, *partial correlation* = .12, *p* = .00) and awareness of consequences (β = .11, *partial correlation* = .14, *p* < .001) were positively associated and significant when controlling for all predictor variables. Biospheric value orientation (β = .03, *partial correlation* = .05 *p* = .18), proenvironmental worldview (β = .00, *partial correlation* = .00 *p* = .83), and the egoistic value orientation (β = .00, *partial correlation* = .10 *p* = .01) were positively associated and nonsignificant; altruism was negatively associated and nonsignificant (β = -.03, *partial correlation* = -.07, *p* = .06).

The influence of age was statistically nonsignificant and primarily positively associated with intention to consume less meat except for participants aged 48-57 (β = -.00, *p* = .75). Most ethnic categories were positively related and nonsignificant, except the mixed category (β = - .01, *partial correlation* = -.05, *p* = .23), which was negatively associated and nonsignificant. Compared to those achieving higher education levels, all categories negatively influenced intention outcomes, and no formal education (β = -.04, *partial correlation* = -.10, *p* = .00) was solely statistically significant.

There was a significant two-way interaction between personal norm and attitude (β = .05, *partial correlation* = .13, *p* < .001), indicating that the effect of personal norm on intention differs depending on the level of attitude. For exercisers, the presence of a climate stewardship personal norm and attitude predicted a statistically significant intention to consume less meat compared to nonexercisers but did not differ between gender. Specifically, as exercisers with proclimate attitude within and above one standard deviation of the mean increased, their moral obligation weakened to predict intention. Exercises below -1 standard deviation of the mean personal norm influence increased in predicting intention across levels of decreasing attitude. The CLES indicates that for a randomly selected exerciser with the moderator of attitude at one standard deviation above the mean and the same level of other predictors, there is a 71.07% chance that exercisers with a higher personal norm will also have a higher intention outcome.

The 48-57 age group (β = -.00, *partial correlation* = .00, p = .94) was negatively and nonsignificantly associated with intention, whereas the remaining age groups were positively and nonsignificantly associated. Mixed participants (β = -.01, *partial correlation* = -.05, *p* = .23) were negatively associated, and all other ethnicities were positively related to intention. Each level of education was negatively associated, and no formal education (β = -.04, *partial correlation* = -.01, *p* = .02) was solely significant.

### VBNTPB Causal Pathway 2 (Attitude & Health Concern) (S13_Table 4)

A significant effect in the VBNTPB causal pathway two was identified when adjoining personal norm, attitude, and health concern as predictors in block five. The VBNTPB casual pathway two differs from the VBN casual pathway in that intention was regressed onto personal norm, attitude, and health concern, increasing the VBNTPB casual pathway two explanation 20.7%. The adjusted R-squared of the model indicates that the model predictors explained 86.4% of the variance in intention, and the model was statistically significant (p < .001).

Personal norm was the most impactful unique significant contributor (β = .39, *partial correlation* = .40, *p* <.001) to predicting intention in the model.

Exercisers’ (b = .602, *partial correlation* = .09, *p* = .02, two-sided) and females’ (b = .247, *partial correlation* = .03, *p* = .36) personal norm, attitude (β = .33, *partial correlation* = .36, *p* < .001), and health concern (β = .16, *partial correlation* = .21, *p* < .001) were more positive and statistically significant associated with intention. Exercisers were significantly different from nonexercises, and females were not significantly different from males. A one-unit increase in exercise status and gender was associated with a 60.2% and 24.7% increase in intention for exercisers and females. This means that the stronger the personal norm, attitude, and health concern of exercisers and females, the stronger their intention to take action in limiting meat consumption to help protect the Earth’s climate. The CLES indicates that a randomly selected exerciser and female with a higher personal norm, attitude, and health concern will also have a higher intention, 52.90% and 51.21% more than nonexercisers or males.

Ascription of responsibility (β = .07, *partial correlation* = .10, *p* = .01) was positively statistically significant, and Biospheric (β = .02, *partial correlation* = .03, *p* =.37) and egoistic (β = .01, *partial correlation* = .03, *p* = .77) value oritentations and ascription of responsibility (β = .07, *partial correlation* = .35, *p* = .45), proenvironmental worldview (β = .00, *partial correlation* = .00, *p* = .82) and awareness of consequences (β = .06, *partial correlation* = .07, *p* = .06) were positively associated and nonsignificant; altruism was negatively associated and nonsignificant (β = -.03, *partial correlation* = -.06, *p* = .09).

Aside from the negatively nonsignificant 48-57 (β = -.00, *partial correlation* = -.00, *p* = .90) age group, the remaining age groups were positively and nonsignificantly associated with intention. The 58-100 age group (β = .02, *partial correlation* = .03, *p* = .36) influenced intention the greatest. Most ethnic groups were positively and statistically nonsignificant; the Mixed (β = - .01, *partial correlation* = -.03, *p* = .36) ethnicity was negatively related and nonsignificant. Compared to the higher education reference group, secondary education (β = .01, partial correlation = *p* = .58) was positive and nonsignificant; the remaining education categories were negative and nonsignificant.

### Two-Way Interaction (Attitude–Health Concern) (S13_Table 4) (S13.1 & 13.2)

There was a significant two-way interaction between attitude and health concern (β = .14, *partial correlation* = .15, *p* < .001), indicating that the effect of attitude on intention differs depending on the level of health concern. Thus, an exerciser’s climate stewardship attitude is moderated by their health concern and is statistically significant in predicting their intention to consume less meat compared to nonexercisers, but does not differ between gender. By this moderation, the influence of attitude on intention was strengthened across all levels of increasing health concern. The CLES indicates that a randomly selected exerciser with health concern as a moderator at one standard deviation above the mean and the same levels of other predictors, there is a 72.87% chance that individuals with a higher personal norm value will also have a higher intention value. No significant two-way interactions were found between personal norm and attitude (β = -.05, *partial correlation* = -.07, *p* = .08) or between personal norm and health concern (β = -.00, *partial correlation* = -.00, *p* = .85).

### Three-Way Interaction (Personal Norm–Attitude–Health Concern) (S13_Table 5) (S13.3)

Exercisers (b = .538, *partial correlation* = .08, *p* = .04) showed a significant three-way interaction across personal norm, attitude, and health concern compared to nonexercisers; females (b = .261, *partial correlation* = .04, *p* = .32) were nonsignificant to males. Accordingly, the interaction between attitude and health concern in the VBNTPB causal pathway two as predictors of intention is influenced negatively by the personal norm predictor of intention. The personal norm–attitude–health concern three-way interaction increased the VBNTPB casual pathway two explanation 21.5%.

The adjusted R-square of the model indicates that the model predictors explained 87.3% of intention variance and were statistically significant (*p* <.001). Personal norms were the most impactful unique significant contributor (β = .41, *partial correlation* = .43, *p* <.001) to predicting intention in the model. Attitude (β = .37, *partial correlation* = .40, *p* <.001) and health concern (β = .22, *partial correlation* = .28, *p* <.001) were positively significant.

A one-unit increase in exercise status and gender was associated with a 53.8% and 26.1% increase in intention for exercisers and females. This means that the stronger the interaction between the personal norm, attitude, and health concern, the stronger the intention of exercisers and females, evaluated by whether participants intended to take action to lower meat consumption to help protect Earth’s climate. The CLES indicates that a randomly selected exerciser and female with a higher personal norm, attitude, and health concern will also have a higher intention, 52.90% and 51.21% more than nonexercisers or males.

Ascription of responsibility (β = .08, *partial correlation* = .12, *p* = .00), awareness of consequences (β = .07, *partial correlation* = .09, *p* = .02), personal norm was positively statistically significant, biospheric (β = .41, *partial correlation* = .43, *p* <.001) and egoistic (β = .01, *partial correlation* = .05, *p* = .23) positively statiscially signfinact. Proenvironmental worldview (β = .01, *partial correlation* = .02, *p* = .58) and were positively associated and nonsignificant; altruism was negatively associated and nonsignificant (β = -.03, *partial correlation* = -.07, *p* = .08).

The three-way interaction of personal norm-attitude-health concern was negatively significant (β = -.19, *partial correlation* = -.25, *p* = <.001). The supporting two-way interactions of personal norm-attitude (β = -.02, partial correlation = -.03, p = .36) were negative and nonsignificant, and then personal norm-health concern (β = .00, partial correlation = .00, p = .99) and attitude—health concern (β = .06, partial correlation = .06, p = .11) were positive and nonsignificant. The three-way interaction indicates that exercisers’ climate stewardship attitude– health concern interaction weakens as personal norm increases. Nonetheless, moderation strength of the attitude-health concern interaction overall is refractive to personal norm, as climate stewardship intention to consume less meat remained statistically significant for exercisers but did not differ between genders. Exercisers’ attitude-health concern interaction values within the mean and above 1 standard deviation influenced personal norm adequately to predict intention significantly. The most pronounced attitude-health concern weakening occurred below -1 standard deviation. Conversely, attitude-health concern was negatively moderated by personal norm. The CLES indicates that a randomly selected exerciser with attitude-health concern as a moderator at one standard deviation above the mean and the same levels of other predictors, there is a 61.32% that individuals with a higher personal norm value will also have a higher intention value.

All age groups were positive and nonsignificant the 58-100 age group was the most impacful in determing intenton outcome of the personal norm-attitude-health concern interaction (β = .03, *partial correlation* = .06, *p* = .10). Most ethnic groups were positively and statistically nonsignificant; the Mixed (β = -.01, *partial correlation* = -.02, *p* = .50) ethnicity was negatively related and nonsignificant. Compared to the higher education reference group, no formal education (β = -.03, *partial correlation* = -.09, *p* = .02) was negative and nonsignificant, whereas the technical/community college and unknown groups were negative and nonsignificant; the secondary level (β = .01, *partial correlation* = -.09, *p* = .03) education was positive and nonsignificant.

### VBNTPB Casual Pathway Three (Attitude, Social Norm, & Personal Norm) (S14)

A significant exerciser (b = .456, β = .02, *partial correlation* = .06, *p* = .05, one-sided) group effect was identified in the VBNTPB model three causal pathway when intention was regressed onto social norm, attitude, and personal norm concurrently in block five and positively associated with intention. Females (b = .498, β = .02, *partial correlation* = .07, *p* = .07) were nonsignificant to males. The VBNTPB model three differs from the VBNTPB one and two causal pathways in that social norm, attitude, and personal norm were concurrent predictors, which increased the explanatory power of the VBNTPB casual pathway three by 20.4%.

The adjusted R-squared value indicates that the model predictors explained 86.1% of variance intention. Social norm (β = .12, *partial correlation* = .21, *p* <.001), attitude (β = .32, *partial correlation* = .33, *p* < .001), and personal norm (β = .38, *partial correlation* = .39, *p* <.001) were positive and significantly associated with intention. Personal norm was the most impactful and unique significant contributor to predicting intention in the model. A one-unit increase in exercise status and gender was associated with a 45.6% and 49.8% increase in intention for exercisers and females. The CLES indicates that a randomly selected exerciser and female with a higher health concern will also have a higher intention, 50.82% and 50.92% more than nonexercisers or males.

Awareness of consequences (β = .11, *partial correlation* = .15, *p* < .001) and Ascription of responsibility (β = .07, *partial correlation* = .10, *p* = .00) were additionally positively and significantly associated with intention. The biospheric (β = .02, *partial correlation* = .02, *p* = .24) and egoistic (β = .00, *partial correlation* = .00, *p* = .99) value orientations were positively associated and nonsignificant when controlling for all predictor variables. Proenvironmental worldview was positively associated and nonsignificant (β = .00, *partial correlation* = .01, *p* = .87); altruism was negatively associated and nonsignificant (β = -.03, *partial correlation* = -.14, *p* = -.06).

The two-way interactions of social norm-personal norm (β = .02, *partial correlation* = .02, *p* = .57) and attitude-personal norm (β = .04, *partial correlation* = -.07, *p* = .07) were positively nonsignificant, and attitude-social norm (β = -.00, *partial correlation* = -.00, *p* = .96). were negatively nonsignificant. **(S14_Table 6)**

All age groups were positively associated, but the association was nonsignificant; individuals aged 58-100 (β = .01, *partial correlation* = .02, *p* = .62) contributed the most influence. Each ethnicity was statistically nonsignificant. Mixed (β = -.01, *partial correlation* = - .04, *p* = .29) participants were negatively associated, and all other ethnicities were positively associated. The technical/community college (β = -.01, *partial correlation* = -.02, *p* = .53), no formal (β = -.03, *partial correlation* = -.08, *p* = .04), and unknown (β = -.01, *partial correlation* = - .04, *p* = .24) education levels were negatively associated with intention; secondary level education (β = .00, *partial correlation* = .00, *p* = .88) was positively associated. The no formal education solely significantly influenced intention.

A three-way interaction of social norm-attitude-personal norm was negatively significant (β = -.19, *partial correlation* = -.25, *p* < .001), but not for exercisers, yet was higher than compared to nonexcericsers. The supporting two-way interactions of personal norm-attitude (β = -.02, partial correlation = -.03, p = .36) were negative and nonsignificant, and then personal norm-health concern (β = .00, partial correlation = .00, p = .99) and attitude-health concern (β = .06, partial correlation = .06, p = .11) were positive and nonsignificant. **(S14_Table 7)**

All age groups were positive and nonsignificantly associated with intention. The Mixed (β = -.01, *partial correlation* = -.03, *p* = .36) ethnicity was negatively related, and those remaining were positively related; all ethnicities were statistically nonsignificant. Each education level was negatively associated and statistically nonsignificant.

### Mediation (S15_Table 8)

The indirect effects of 5000 bootstrapping analyses supported mediation within the VBN causal pathway and its VBNTPB causal pathway iterations, as indicated by the bias-corrected and accelerated 95% confidence intervals, which did not contain zero, indicating statistical significance (Hayes et al., 2011; Hoeksma et al., 2017). The positive and significant mediating relations support the VBN causal pathway explanation of intention originating from environmental orientation values. Proenvironmental worldview mediated the relationship between biospheric, altruistic, and egoistic value orientations and awareness of consequences, the awareness of consequences between proenvironmental worldview and ascription of responsibility, the ascription of responsibility between awareness of consequences and personal norm, personal norm between ascription of responsibility and intention.

The inclusion of rational evaluation variables in the context of self-interest influenced personal norm, resulting in the VBNTPB causal pathways’ significance for intention. In contrast, the VBN causal pathway was not significant for exercises without these variables. Personal norm remained as a mediator in the VBNTPB pathways, and its indirect effect was reduced, supporting the increased explanatory power by the self-interest variables.

### Diet Modification

A chi-square test of association was conducted to assess the relationship between exercisers and nonexercisers and diet modification to consume less meat to benefit Earth’s climate. Of the exerciser (exposed) group, 115 participants attested to changing their meat consumption behavior to reduce unnatural climate change compared to 76 in the nonexerciser (nonexposed) group. One hundred fifty-five exercisers and 209 nonexercisers did not alter their meat consumption. The association between exercise status and diet modification to reduce unnatural climate change was statistically significant, *X^2^*(1, *N* = 555) = 15.58, *p* <.001. The odds ratio magnitude of association was 2.04 indicating exercisers were 100% more likely than nonexercisers modify their meat consumption behavior to reduce unnatural climate change (*OR* = 2.04, 95% *CI* [1.42, 2.91]).

**Fig 3.**
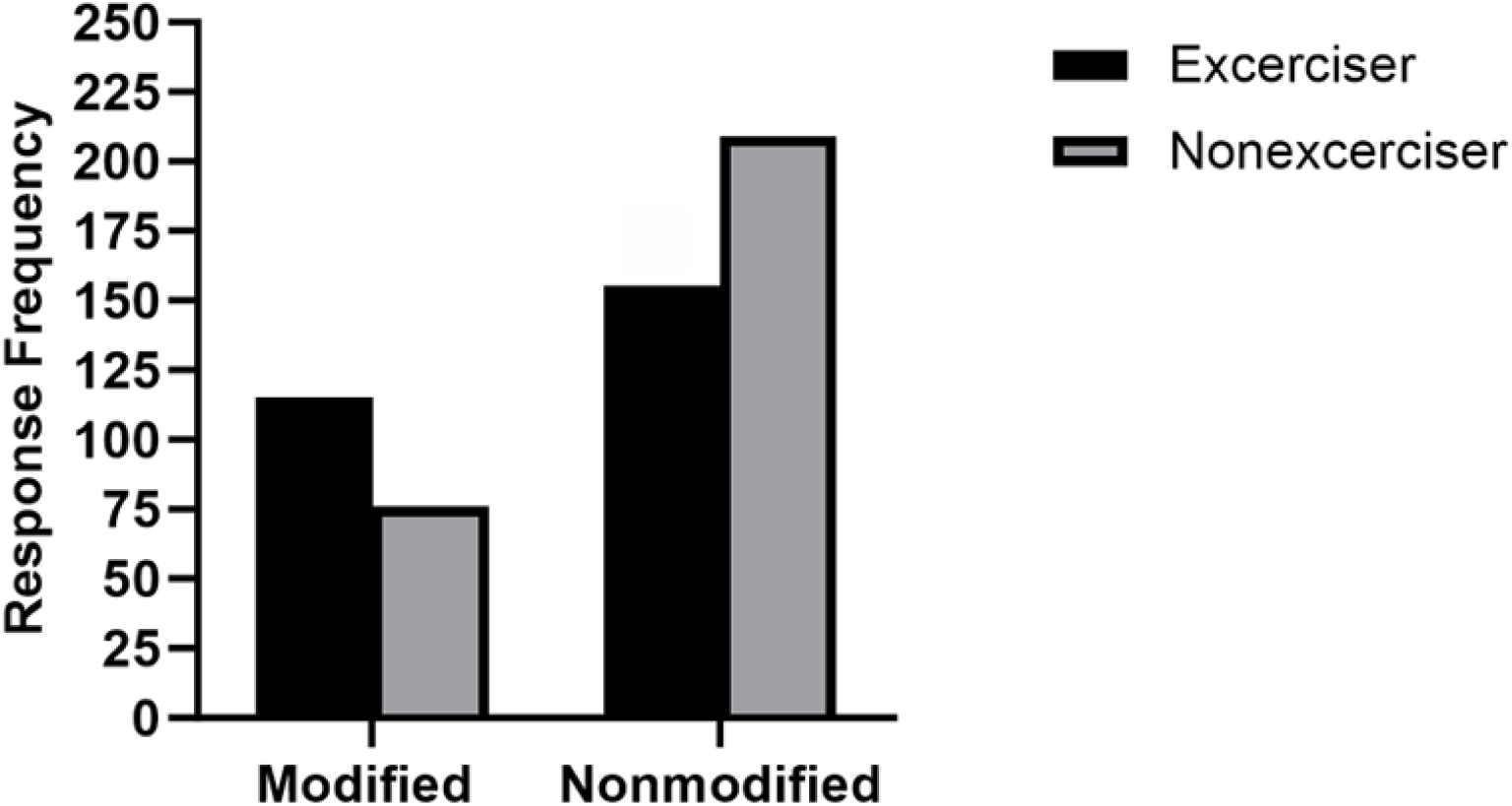
Diet Modification to Reduce Unnatural Climate Change and Exercise Status.

## Discussion

Human compliance in following a low-meat diet can help prevent unwanted climate change, primarily described as global warming. Physically active individuals inherently complete environmentally friendly behaviors—as physical activity increases, so do physically active individuals’ willingness and beliefs in performing eco-friendly behaviors (Cunningham et al., 2020; Teixeira et al., 2023; Thormann & Wicker, 2021; Thormann & Wicker, 2022; Wicker & Thormann, 2022). Relevant studies have not determined what behavioral antecedents in exercisers may be leveraged or within a causal pathway to conclude an individual’s intention in carrying out climate-friendly behavior. A study was conducted to determine if exercise status is associated with environmental value orientations, a proenvironmental worldview, willingness, and behavior to limit human meat consumption to protect Earth’s climate.

Of the 12 psychosocial constructs examined, exercisers’ biospheric, altruistic, and egoistic value orientations, proenvironmental worldview, awareness of consequences, ascription of responsibility, attitude, health concern, personal norm, social norm, concluding with intention were statistically significant compared to nonexercisers when arranged in a causal pathway, reflective of the value belief norm theory (VBN), theory of planned behavior (TPB) models and a health concern variable to form an integrated VBNTPB model. Exercisers sequentially predicted each VBNTPB causal pathway variable more strongly and significantly than nonexercisers, leading to a positive and significant intention to consume less meat to protect the Earth’s climate.

The group significance of exercisers and utility of the combined causal pathway variables in evaluating the promotion of environmental citizenship is robust, as indicated by the large common language effect sizes exceeding 50%. These effect sizes indicate that more physically active individuals and females will exhibit higher outcomes for each predictor variable than less physically active individuals, and the VBNTPB causal pathways explain more variance than the VBN causal pathway. Moreover, a measurement of exercisers’ current behavior found they followed a low-meat diet 100% more than nonexercisers to limit unwanted climate change.

The VBNTPB model was formed by integrating the TPB and health concern variables at the level of the VBN personal norm prediction of intention. Statistically significant intention outcomes were confirmed in three distinct VBNTPB causal pathways. In VBNTPB causal pathway one, the intention variable was regressed onto the variables of personal norm and attitude. In the second VBNTPB causal pathway, intention was regressed onto attitude, health concern, and personal norm. Intention was regressed onto social norm, attitude, and personal norm in the third VBNTPB causal pathway. The variables of attitude, health concern, social norm, and personal norm were the most proximal predictors of intention. The VBN model outcomes also predicted each causal pathway variable positively and significantly, except for exercisers’ personal norm, which predicted intention nonsignificantly compared to nonexercisers.

The results indicate that the VBNTPB model causal pathways successfully explained how more physically active individuals formulate intentions to practice climate stewardship by modifying their diets to consume less meat. As each causal pathway variable was added, the VBNTPB causal pathways adjusted R-square explanatory power increased, explaining between 85.4%–87.3% of intention variance, confirming the model’s design compared to the VBN casual pathway, which explained 83.2%. Thus, individuals who exercise to maintain optimal health are more inclined towards biospheric, altruistic, and egoistic value orientations that shape a proenvironmental worldview, where they recognize that human actions negatively impact the environment. This recognition influences their behavior-specific beliefs and norms, leading to a reduction in human meat consumption to benefit Earth’s climate and intentions to carry out future climate stewardship.

The VBNTPB causal pathway one identified that exercisers predicted climate stewardship intention significantly different than nonexercisers. The positive and significant moderation of personal norm by attitude and significant intention outcome, compared to the VBN causal pathway, indicates that attitude is a driver of personal norm development, such that the sense of not feeling morally obligated is weakened. Exercisers’ attitudes or settled ways of thinking and feeling influence their personal norm, and their attitude reflects their moral obligation to limit meat consumption to benefit the Earth’s climate. Attitude presence indicates reasoned decision-making that is favorable for executing an intention (Carfora, 2023; Chen, 2024; Miller et al., 2022; Seffen & Dohle, 2023). Chen’s (2024) results further support this study’s findings of how the rational decision-making influence of attitude moderates a nonsignificant proenvironmental intention to a significant proenvironmental intention outcome.

Various mixed integrated VBN/TPB models have been successful in significantly regressing proenvironmental intention onto proenvironmental attitude (Carfora et al., 2021; Fauzi et al., 2024; Han, 2015; Laksmana & Hendriana, 2023).

In further support of exercisers’ beneficial attitude towards promoting climate stewardship, the health concern VBNTPB causal pathway two variable moderated attitude positively significantly, and in combination with the personal norm variable, positively and significantly predicted intention. Personal norm was negatively impacted in each two-way interaction with attitude and health concern, yet nonsignificant, indicating that the positive, significant intention outcomes derive from the interaction of attitude and health concern and relieve the inadequacy of personal norm. As such, the intention to limit meat consumption for climate stewardship does not originate solely from a sense of moral obligation, but also from exercisers’ health concerns for their health and that of others, and their attitude, supported by health concerns.

The nonsignificant exerciser personal norm-health concern interaction is not entirely surprising, given the controversy of meat’s impact on health, views of not consuming meat as unhealthy, the psychological pleasure, and sociocultural expectations of meat consumption (Sanchez-Sabate et al., 2019). The health concern construct indicates if participants considered meat consumption potentially harmful to sustaining good health, the climate, and whether the two were interdependent. Meat is known to cause illness (Battaglia et al., 2015; Neff et al., 2018). Therefore, it is logical to conclude that those concerned with good health, especially exercisers pursuing optimal health, would feel obligated to benefit human health and Earth’s climate by intentionally limiting meat consumption. The results of this study confirm that idea.

Despite exercisers’ accomplishments of routine physical activity to maintain optimal health, their limited personal norm reasoning to carry out climate stewardship was not solely benefited by health concern.

A significant three-way interaction between attitude, health concern, and personal norm was also negative, supporting the same conclusion and that the relationship between attitude and health concern varies depending on the level of an exerciser’s personal norm. In other words, the enhancing attitude-health concern interaction with personal norm when predicting exercisers intention is weakened as personal norm levels increase. Exercises with the lowest personal norm, 1 standard deviation below the mean, were largely refractory to the enhancing attitude-health concern interaction. Supportively, the attitude-health concern interaction was reversed when moderated by personal norm.

Carfora (2021) found that personal norm significantly predicted the intention to purchase natural foods to benefit the environment. An included trust variable predicted the same, implying concern for food safety and, consequently, health. Lai et al. (2020) found that their VBNTPB model identified significant effects on reducing meat consumption intention related to protecting the environment and personal health, which were predicted by personal norm and health concern variables, respectively. Health messages led to stronger intentions than environmental messages to reduce meat consumption for the betterment of climate change (Tallie et al. 2022). The results of this study and those aforementioned indicate that the self-interest rational motives of health concern and those of TPB, help individuals rationalize their personal norm moral motive to support environmentalism.

The nonsignificant predictive interaction between personal norm and health concern strengthens the presence of biospheric, altruistic, and egoistic self-identity environmental value orientations. Exerciser value orientations positively and significantly predicted a proenvironmental worldview, which was further transmitted through the causal pathways to influence the origination of proenvironmental beliefs and norms. The results indicate these values are inherent within exercisers as the overarching drivers for the causal pathway significant intention to modify meat consumption to protect Earth’s climate. Exercisers’ biospheric, altruistic, and egoistic environmental value orientations support their care for the environment, welfare for others, and self-concern for themselves. There is no trade-off between biospheric and altruistic value orientations compared to egotism (Lou et al., 2024; de Groot & Steg, 2010; Schultz et al., 2005). The self-interest of exerciser egotism aligns with caring for the environment and humanity while sustaining indoor and outdoor exercise for themselves through atmospheric environmentalism (de Groot & Steg, 2010; Schultz et al., 2005; Wallace et al., 2019).

Additionally, human values vary according to geographical surroundings (Chen et al., 2020; Rentfrow, 2010). Differences in lifestyle and economics may strengthen or weaken values. The participants of this study resided across the United States, yet they were mainly in the South. The lack of regional variation could moderate the VBN value orientations due to selective migration, ecological influence, and social influence. Concisely, localities’ physical and social features can affect feelings, psyche, and behaviors (Rentfrow, 2010).

The VBNTPB casual pathway three showed no significant exerciser two-way or three-way interactions between social norm, attitude, and personal norm for intention; however, the main effects for each variable when predicting intention were positive and significant for exercisers compared to nonexercisers. This means that the positive and significant influence of social norm, attitude, and personal norm is consistent individually to predict intention regardless of the levels of the three variables. The result adds that significant social pressure exists among exercisers to curb meat consumption for the benefit of climate stewardship, and increases the variance explanation of intention compared to the VBN causal pathway.

Variation has been shown to exist in social norms to influence climate-friendly behavior in TPB models (Bradley et al., 2024; Yoon et al., 2010; Masud et al., 2016). Braksiek et al. (2021) identified a significant TPB effect of sports club members’ social norms to act environmentally friendly. Chen’s (2024) VBN/TPB merged model found no significant effect of social norm prediction on intention to purchase suboptimal foods suitable for consumption to benefit the environment. Aside from Braksiek et al. (2021), the preceding study populations were not physically active individuals; differences in geographical location and the context of performing the environmentally friendly behavior may affect the outcomes. Çoker et al. (2022) found that the significant influence of social norms on choosing foods to benefit health and the environment was dependent on whether individuals of social norm groups consume food together or not. Whether exercisers in this study, as a social norm group, consumed food together with other exercisers may affect exercisers’ social norm, attitude, and personal norm interactions.

Although Stern (1999) postulated personal norm as the most pivotal variable in activating proenvironmental behavior, a significant positive result was not found when the intention variable was regressed onto personal norm in the VBN causal pathway. Personal norm of the VBN casual pathway explained 51.8% of intention variance, and personal norm predictive power was in line with and outperformed related studies, ranging from 5‒55.7% (Aguilar-Luzon et al., 2012; Fauzi et al., 2024; Hiratsuka et al., 2018; Hoeksma et al., 2017; Hong et al., 2024; Steg et al., 2005; Yang et al., 2024; Zhang et al., 2020) when evaluating proenvironmental intention. The nonsignificant predictive personal norm to proenvironmental diet alteration intention, without rational evaluation variables, supports previous conclusions that personal norm predictive power dampens by more effortful and costly tasks to facilitate environmentalism (Arya & Kumar, 2023; Kácha & Linden, 2021; Loo et al., 2023; Niu et al., 2023; Steg et al., 2005; Stern, 2000; Wang et al., 2024).

The VBNTPB significant casual pathways personal norm variance was reduced and ranged from 16% to 20.4% amongst the three significant causal pathways and relevant interactions. Similar VBN and TPB merged model results were found for personal norm prediction of proenvironmental intention to purchase more costly green computers and increased effort to reduce food waste through consumption habits (Chen, 2024 & Loo et al., 2023).

The requirement of rational evaluation variables within the VBN causal pathway to significantly predict intention suggests that, in the context of altering a meat diet to promote climate stewardship, the sense of morally right behavior, although impactful, is not sufficient. In each VBNTPB causal pathway, personal norm unique variance was reduced in conjunction with attitude, health concern, and social norm, yet personal norm remained the greatest unique predictor, followed by attitude and then health concern or social norm. Rationalizing that personal norm predisposes exercisers to formulate positive intentions. Regarding the value action gap in this context, individuals who possess deep-seated morality do not always behave accordingly, but their disposition may change depending on their evaluation of favorable or unfavorable outcomes, health concerns, and social pressure.

Exercisers’ perceived behavioral control to reduce their meat consumption to mitigate climate change was not statistically significant compared to nonexercisers. The finding was applicable across all iterations of personal norm and PBC in conjunction with attitude, health concern, or social norm. Braksiek et al. (2021) identified a TPB significant effect of sports club members’ perceived behavioral control to carry out environmentally friendly behavior, although the level of physical activity was not quantified; participants were classified only by their sport of choice. Other VBN/TPB merged models PBC predicted significant proenvironmental intentions to reside in green hotels and support environmentally-conscious food choices (Chen, 2024; Fauzi et al., 2024; Han, 2015). Consistent with this study, Loo’s (2024) merged VBN/TPB model, and the increased cost characteristic of purchasing green computers PBC prediction of behavioral intention was nonsignificant. Cost and effort are relevant factors to transition and sustain a low-meat diet.

The intention of exercisers to consume less meat to limit unwanted climate change is higher than that of nonexercisers—indicating an assumption of planning and deliberate decisions to reduce future meat consumption to prevent climate degradation. Intention has been reported as a significant outcome when examining environmentally friendly behavior to prevent unwanted climate change. Thormann and Wicker (2022) identified that as environmental friendliness and sports club members’ practice hours increased, so did their willingness to pay a carbon tax to reduce global warming. Braksiek et al. (2021) found that the intention of sports club members to act environmentally friendly was statistically significant between constructs within the Theory of Planned Behavior (TPB) model. Both studies and this study’s findings support the existence of internal traits in routinely physically active individuals that enable them to carry out climate-friendly behavior.

Exercisers’ current behavior of reducing their meat consumption to limit unwanted climate change was more frequent than that of nonexercisers. Exercisers modified their meat consumption habits to benefit Earth’s climate 100% more than nonexercisers. Teixeria et al. (2022) measured the current status of physical activity and proenvironmental behavior, such as recycling, green travel requiring physical movement, and encouraging others to practice environmentally sustainable practices. In their study, physical activity correlated with a range of proenvironmental behaviors.

The misalignment of a significant diet alteration odds ratio and a nonsignificant PBC indicates that exercisers’ control beliefs were not supportive of diet alteration engagement. The outcome suggests that social norm, attitude, health concern and personal norm more heavily influenced exercisers’ climate stewardship meat reduction behavior.

Sociodemographic trends were identified. Compared to the 18-27 reference age group, all other age groups positively influenced the VBN causal pathway outcomes, and the beta coefficients routinely increased concurrently with age. In contrast, all age groups negatively impacted the prediction of intention. Similar trends were observed in the VBNTPB models 1-3. Although the statistical significance between age groups was few, the data trends suggest that a stronger commitment to environmentalism is linked to increasing age.

Aside from the negative influence of Mixed ethnicity throughout each sequential causal pathway analysis, with Whites as the reference category, the categorization of Asian, Black, and Other ethnicities had a positive impact on the VBN pathway outcomes. The same was observed for VBNTPB models 1-3. Statistical significance was primarily not found. The findings suggest that individuals who identify with more than one ethnicity have different cultural experiences that affect their environmental friendliness.

Lower education levels, including technical/community college, secondary, no formal education, or unknown education, primarily negatively influenced the outcomes of the VBN and VBNTPB causal pathways compared to higher education as the reference group, which encompassed undergraduate and graduate levels. The outcomes imply that as individuals become more knowledgeable through academic learning, their exposure helps create an environmentalism mindset.

This study targeted a balanced assessment between exercisers and nonexercisers by geography and age, supporting better external validation than routinely cited throughout the literature when measuring environmental friendliness and exercise status (Braksiek et al., 2023; Teixeria et al., 2022; Thormann & Wicker, 2021; Thormann & Wicker, 2022; Wicker & Thormann, 2022).

### Limitations

Self-report bias is a potential limitation since participants self-qualified their exercise habits to create two groups of exercisers and nonexercisers. If participants under- or overestimated their exercise level, it limits the ability to conclude how exercisers’ and nonexercisers’ environmentalism differ and their willingness to modify their diets to benefit the environment. The collection of untrustworthy responses was minimized since each participant completed a lifestyle profile attesting to their exercise habits well before study participation.

Moreover, each participant’s exercise habits were confirmed when entering the survey. Those whose responses did not agree with their lifestyle profile were omitted.

A data analysis limitation exists because the performance of the CFA could not produce a clearly defined matrix table of loaded factors. The CFA analysis implies uncertainty in whether the survey questions measured the ideas intended to be measured. Reducing the number of survey constructs may be helpful (Ross & Zaidi, 2019). Several survey iterations were completed to minimize obtaining an unclear matrix table. Although the data satisfied requirements of multiconlinearity, because the survey questions were highly related, producing a verifiable matrix table of loaded factors is a known barrier (Muller-Schneider, 2022; Watkins, 2018).

A representative sample of the United States population was targeted to complete the study in Prolific’s crowdsourcing database. The representative profile could vary from the true demographics, thereby producing responses that were not representative of the general population. In reviewing the 2023 United States census ethnic data, the population groups targeted varied by no more than .9% (U.S. Census Bureau, n.d.). Other demographic data, such as U.S. region of residence, education level, and exercise status, were unavailable. Kraemer et al. (2017) reported that demographic differences might exist when relying on crowdsourced participants.

Because survey deployment was via the internet, a location threat is possible. The survey location settings of the participants are unknown to the researcher. Given the timeliness of survey completion and the disqualification of speedy survey takers, it is unlikely that any unusual systematic circumstances affected participant completion.

Instrument decay may have occurred during the completion of the 63 and 64-item nonexerciser and exerciser surveys, leading to response fatigue. As mentioned, participants completed the survey items promptly, indicating clarity of participant understanding. Durations beyond three standard deviations of the average time were omitted. The completion times between the two populations varied by .9%, suggesting that response fatigue did not affect study results.

### Recommendations

Despite evidence of survey length adequacy, shorter surveys are generally considered more desirable than longer instruments. Such an alteration can improve participants’ focus on what constructs are evaluated and is particularly important when evaluating closely related multiple subscales, as in this research. Condensing the survey and correlating outcomes between multiple survey iterations is a possible future strategy.

Further survey development is needed to produce a verifiable CFA matrix table, so long as the observable indicators per construct target different related viewpoints of each construct. Many surveys dependent on latent variables do not. A researcher may consider drafting more straightforwardly written survey items. Because of the survey length, the researcher cautiously constructed items that would not lead participants. Given that meat consumption, the production of GhG, and unwanted climate change are factual, removing the conditional phrases can simplify survey item wording. It may improve the production of a verifiable CFA matrix table.

Measuring observable data on current behavior is prudent to advance understanding of physical activity as a mediator of environmentally friendly behavior. A limitation exists between what individuals report and their actual actions. Since exercise status is the qualification for study participation and grouping, documenting exercise status without potential participants’ knowledge can avoid self-report bias when qualifying participants. The same strategy can offset measurement errors in diet behavior. A future direction may consider measuring individual meat consumption first to correlate with environmental friendliness scores afterward. This study relied on previously reported physical activity status unrelated to the study and confirmed that status and reasoning for physical activity to benefit health prior to survey admittance, concluding the removal of 40.4% participants from the exerciser and nonexerciser study groups. A short-term covert longitudinal study could fully satisfy any concern of self-bias.

To enhance proenvironmental outreach, employers should develop and incorporate sustainability goals at their company level and evaluate prospective employee environmental stewardship mindsets as an employment qualification (Ones et al., 2015). Embedding environmentally friendly behavior must be structured as to what is acceptable, that is, to be responsible and contribute to a more significant cause. Environmental consciousness will become enhanced through routine daily behavior and increased information provisioning through employment channels. Mandates and training at worksites can shift mindsets to promote environmental friendliness.

### Conclusions

An analysis of exercisers’ environmental views and willingness to reduce meat consumption to prevent unwanted climate change was completed using an aggregated model consisting of the VBN theory, NEP scale, a health concern construct, and the theory of planned behavior. The outcomes indicate that individuals who complete the physical activity of exercise at the level recommended by the U.S. Department of Health and Human Services (2024) and World Health Organization (2022) guidelines—to sustain the best health—possess values, proenvironmental beliefs, norms, and intentions that influence their willingness to act more environmentally friendly than less physically active individuals (U.S. Department of Health and Human Services, 2024; WHO, 2022).

This study is the first to reveal three salient findings regarding the relationship between physical activity and environmentalism. At the individual level, this study identified a significant association between individuals who exercise regularly, biospheric, altruistic, and egoistic value orientations, awareness of consequences, ascription of responsibility, personal norm, attitude, health concern, social norm, and intention to protect the climate from degradation caused by GhG released from animal rearing to produce meat for human consumption. Secondly, individuals who exercise regularly are twice as likely as nonexercisers to practice climate stewardship by consuming less meat. Thirdly, rational evaluation can be a necessity for influencing a moral disposition to act proenvironmentally. In this study, exercisers within the

United States were willing to consume less meat voluntarily to benefit the climate via an evaluation of self-interest.

Personal norm, attitude, health concern, and social norm were the most proximal indicators of behavior change, indicating that in exercisers, they are more influential than perceived behavioral control in promoting reduced meat consumption to protect the climate. The constructs indicate potential intervention points in environmental value orientations, beliefs, and norms. Since nearly half of U.S. adults meet at least one of the U.S. Health and Human Services physical activity recommendations, the Earth’s climate may benefit by leveraging exercisers to promote less consumption of meat.

The transference of exercisers’ inherent personal norm and self-interest to those less physically active may be accomplished by environmental activism to enact legislative policies promoting less meat consumption, such as a meat tax, meat substitutes, and plant-based dieting rebate programs, health care incentives, and restrictions on meat availability and its consumption. Future studies should be conducted using similar integrated behavior theories, within and beyond the United States, to continue support of the association between physical status and environmentalism, and to identify other inherent traits that may be useful targets of social change advocacy. Particularly, how rational evaluation in the context of self-interest can enhance personal norms to accomplish proenvironmental intentions and behavior. Identifying other preexisting groups with such inherent traits may create a foundation for more widespread advocacy to limit meat consumption for climate benefits and ensure the continued existence of humankind.

## Data Availability

Some data is available and contained within the submission documents.

## Acknowledgments

The author thanks the professionally accredited statistician, who provided expert statistical assistance to complete the study.

