## Supplemental File 9 for "Determinants of Exerciser Environmentalism to Lower Meat Consumption in the Practice of Climate Stewardship"

**S9 File**

**Descriptive Statistics for Construct Groups**

| Constructs | Exerciser | | | | Nonexerciser | | |
|---|---|---|---|---|---|---|---|
| | *n* | *SD* | *Mdn* | *IQR* | *SD* | *Mdn* | *IQR* |
| Biospheric | 4 | 3.16 | 25.00 | 4.00 | 2.86 | 24.00 | 4.00 |
| Altruistic | 5 | 3.64 | 31.00 | 4.00 | 3.63 | 31.00 | 5.00 |
| Egoistic | 3 | 3.75 | 8.00 | 5.00 | 3.45 | 8.00 | 5.00 |
| Proenvironmental view | 6 | 7.06 | 31.00 | 9.00 | 6.62 | 32.50 | 9.50 |
| Awareness of consequences | 5 | 7.27 | 27.00 | 8.00 | 7.29 | 26.00 | 8.50 |
| Ascription of responsibility | 5 | 5.99 | 22.00 | 8.00 | 6.08 | 23.00 | 7.00 |
| Attitude | 5 | 6.08 | 22.00 | 9.00 | 6.72 | 21.00 | 10.00 |
| Personal norms | 5 | 7.76 | 22.00 | 11.00 | 7.56 | 20.00 | 12.00 |
| Perceived behavioral control | 4 | 5.28 | 20.00 | 7.00 | 5.53 | 18.00 | 8.00 |
| Social norms | 4 | 4.65 | 14.50 | 6.00 | 4.57 | 13.00 | 6.00 |
| Health concern | 4 | 5.39 | 20.00 | 6.00 | 4.00 | 19.00 | 7.00 |
| Intention | 5 | 8.65 | 20.00 | 6.00 | 8.56 | 22.00 | 15.00 |
