## Supplemental File 10 for "Determinants of Exerciser Environmentalism to Lower Meat Consumption in the Practice of Climate Stewardship"

**S10 File**

**Tukey Fence Exerciser and Nonexcerciser Construct Response Percent Outliers**

| | Exercisers $n$ = 300 | Nonexcercisers $n$ = 300 |
|---|---|---|
| | Percent Outliers | |
| Construct | | |
| Biospheric (4 items) | 2 | 0.67 |
| Altruistic (5 items) | 3.33 | 1.67 |
| Egoistic (3 items) | 1.67 | 1.33 |
| Proenvironmental View (6 items) | 2 | 1 |
| Awareness of Consequences (5 items) | 4.33 | 4 |
| Ascription of Responsibility (5 items) | 0.33 | 3.67 |
| Attitude (5 items) | 0 | 0 |
| Personal Norms (5 items) | 0 | 0 |
| Perceived Behavior Control (4 items) | 1.33 | 0 |
| Social Norms (4 items) | 1.35 | 0 |
| Health Concern (4 items) | 4.33 | 2.13 |
| Intention (5 items) | 0 | 0 |
