## Supplemental File 11_Table 2 for "Determinants of Exerciser Environmentalism to Lower Meat Consumption in the Practice of Climate Stewardship"

**Table 2**
Results of regression multiple regression analyses of the VBN causal pathway to reduce meat consumption for climate stewardship.

| | b | β | *t* | *p* | S.E. | Partial correlation | 95% CI $R^2$ | Adj $R^2$ | df | Sig *F* change | Δ $R^2$ |
|---|---|---|---|---|---|---|---|---|---|---|---|
| *DV: Intention* | | | | | | | [.80-.85] | .83 | 1, 578 | <.001 | .175 |
| Group | .318 | .01 | 1.04 | .29 | .30 | .04 | | | | | |
| Gender | .457 | .02 | 1.51 | .13 | .30 | .06 | | | | | |
| Personal norm | | .69 | 24.96 | <.001 | .03 | .47 | | | | | |
| Ascription of responsibility | | .15 | 5.03 | <.001 | .04 | .12 | | | | | |
| Awareness of consequences | | .11 | 3.37 | <.001 | .03 | .12 | | | | | |
| Proenvironmental worldview | | .01 | .42 | .67 | .03 | .00 | | | | | |
| Biospheric | | .02 | 1.16 | .24 | .07 | .04 | | | | | |
| Altruistic | | -.03 | -1.58 | .11 | .06 | -.06 | | | | | |
| Egoistic | | .01 | .79 | .42 | .04 | .03 | | | | | |
| 28-37 | | -.01 | -.86 | .39 | .47 | -.03 | | | | | |
| 38-47 | | -.01 | -.70 | .48 | .48 | -.02 | | | | | |
| 48-57 | | -.02 | -1.07 | .28 | .48 | -.04 | | | | | |
| 58-100 | | -.01 | -.56 | .57 | .51 | -.02 | | | | | |
| Asian | | .00 | .27 | .78 | .75 | .01 | | | | | |
| Black | | .02 | 1.34 | .17 | .54 | .06 | | | | | |
| Mixed | | -.01 | -.96 | .33 | .67 | -.04 | | | | | |
| Other | | .00 | .41 | .92 | .74 | .01 | | | | | |
| Technical/Community College | | -.02 | -1.39 | .16 | .40 | -.05 | | | | | |
| Secondary | | -.01 | -.84 | .40 | .35 | -.03 | | | | | |
| No formal | | -.04 | -2.62 | .00 | 1.48 | -.10 | | | | | |
| Unknown | | -.01 | -1.10 | .26 | 1.47 | -.04 | | | | | |
| *DV: Personal norm* | | | | | | | [.60-.69] | .65 | 1, 579 | <.001 | .073 |
| Group | 1.06 | .06 | 2.44 | .01 | .43 | .10 | | | | | |
| Gender | 1.14 | .06 | 2.65 | .00 | .43 | .11 | | | | | |
| Ascription of responsibility | | .44 | 11.23 | <.001 | .05 | .42 | | | | | |
| Awareness of consequences | | .30 | 6.74 | <.001 | .05 | .27 | | | | | |
| Proenvironmental worldview | | .02 | .65 | .51 | .05 | .02 | | | | | |
| Biospheric | | .14 | 4.33 | <.001 | .09 | .17 | | | | | |
| Altruistic | | -.04 | -1.25 | .20 | .09 | -.05 | | | | | |
| Egoistic | | .07 | 2.90 | .00 | .06 | .12 | | | | | |
| 28-37 | | -.00 | -.05 | .95 | .68 | -.00 | | | | | |
| 38-47 | | .00 | .12 | .90 | .69 | .00 | | | | | |
| 48-57 | | .04 | 1.22 | .22 | .70 | .05 | | | | | |
| 58-100 | | .05 | 1.55 | .12 | .73 | .06 | | | | | |
| Asian | | -.00 | -.30 | .76 | 1.08 | -.01 | | | | | |
| Black | | .04 | 1.61 | .10 | .78 | ..06 | | | | | |
| Mixed | | -.02 | -.97 | .33 | .96 | -.04 | | | | | |
| Other | | .02 | .98 | .32 | 1.06 | .04 | | | | | |
| Technical/Community College | | -.00 | -.06 | .94 | .58 | -.00 | | | | | |
| Secondary | | .00 | .25 | .80 | .51 | .01 | | | | | |

| | | | | | | | | | | | |
|---|---|---|---|---|---|---|---|---|---|---|---|
| No formal | | -.04 | -1.88 | .06 | 2.13 | -.07 | | | | | |
| Unknown | | -.03 | -1.47 | .14 | 2.12 | -.06 | | | | | |
| *DV: Ascription of responsibility* | | | | | | | [.52-.62] | .57 | 1, 580 | <.001 | .150 |
| Group | 1.24 | .07 | 2.58 | .01 | .48 | .10 | | | | | |
| Gender | 1.42 | .08 | 2.99 | .00 | .47 | .12 | | | | | |
| Awareness of consequences | | .59 | 14.5 | <.001 | .04 | .51 | | | | | |
| Proenvironmental worldview | | .08 | 1.82 | .06 | .05 | .07 | | | | | |
| Biospheric | | .15 | 4.06 | <.001 | .10 | .16 | | | | | |
| Altruistic | | -.01 | -.34 | .72 | .10 | -.01 | | | | | |
| Egoistic | | .11 | 4.25 | <.001 | .06 | .17 | | | | | |
| 28-37 | | .02 | .80 | .42 | .75 | .03 | | | | | |
| 38-47 | | .04 | 1.13 | .25 | .76 | .04 | | | | | |
| 48-57 | | .08 | 2.32 | .02 | .76 | .09 | | | | | |
| 58-100 | | .10 | 2.89 | .00 | .80 | .11 | | | | | |
| Asian | | .02 | .95 | .34 | 1.18 | .03 | | | | | |
| Black | | .06 | 2.16 | .03 | .86 | .08 | | | | | |
| Mixed | | -.03 | -1.08 | .27 | 1.06 | -.04 | | | | | |
| Other | | .02 | .96 | .33 | 1.17 | .04 | | | | | |
| Technical/Community College | | -.01 | .57 | .56 | .64 | .02 | | | | | |
| Secondary | | -.01 | -.03 | .96 | .56 | -.00 | | | | | |
| No formal | | -.03 | -1.32 | .19 | 2.34 | -.05 | | | | | |
| Unknown | | -.04 | -1.64 | .10 | 2.33 | -.06 | | | | | |
| *DV: Awareness of consequences* | | | | | | | [.36-.48] | .42 | 1, 581 | <.001 | .118 |
| Group | 1.73 | .10 | 3.09 | .00 | .56 | .12 | | | | | |
| Gender | .66 | .03 | 1.19 | .23 | .55 | .05 | | | | | |
| Proenvironmental worldview | | .46 | 11.03 | <.001 | .05 | .41 | | | | | |
| Biospheric | | .18 | 4.22 | <.001 | .12 | .17 | | | | | |
| Altruistic | | .06 | 1.53 | .12 | .12 | .06 | | | | | |
| Egoistic | | .11 | 3.51 | <.001 | .07 | .14 | | | | | |
| 28-37 | | .00 | .04 | .96 | .88 | .00 | | | | | |
| 38-47 | | .00 | .09 | .92 | .89 | .00 | | | | | |
| 48-57 | | .01 | .36 | .71 | .88 | .01 | | | | | |
| 58-100 | | .04 | .94 | .34 | .93 | .03 | | | | | |
| Asian | | .04 | .1.32 | .18 | 1.37 | .05 | | | | | |
| Black | | .05 | .1.80 | .07 | 1.01 | .07 | | | | | |
| Mixed | | -.05 | -1.67 | .09 | 1.24 | -.06 | | | | | |
| Other | | .04 | 1.23 | .21 | 1.37 | .05 | | | | | |
| Technical/Community College | | -.03 | -1.07 | .28 | .74 | -.04 | | | | | |
| Secondary | | -.08 | -2.30 | .02 | .64 | -.09 | | | | | |
| No formal | | -.03 | -1.11 | .26 | 2.74 | -.04 | | | | | |
| Unknown | | -.05 | -1.59 | .11 | 2.73 | -.06 | | | | | |
| *DV: Proenvironmental worldview* | | | | | | | [.24-.36] | .30 | 17, 582 | <.001 | .322 |
| Group | 1.22 | .07 | 2.00 | .04 | .61 | .08 | | | | | |

| | | | | | | |
|---|---|---|---|---|---|---|
| Gender | 1.43 | .08 | 2.39 | .01 | .60 | .09 |
| Biospheric | | .40 | 9.42 | <.001 | .12 | .36 |
| Altruistic | | .15 | 3.65 | <.001 | .13 | .15 |
| Egoistic | | .07 | 2.00 | .04 | .08 | .08 |
| 28-37 | | .00 | .19 | .84 | .96 | .00 |
| 38-47 | | .02 | .46 | .64 | .98 | .01 |
| 48-57 | | .02 | .52 | .60 | .97 | .02 |
| 58-100 | | .02 | .43 | .66 | 1.02 | .01 |
| Asian | | .07 | 2.06 | .03 | 1.51 | .08 |
| Black | | .04 | 1.37 | .17 | 1.11 | .05 |
| Mixed | | -.04 | -1.21 | .22 | 1.36 | -.05 |
| Other | | .05 | 1.50 | .13 | 1.51 | .06 |
| Technical/Community College | | -.02 | -.69 | .48 | .81 | -.02 |
| Secondary | | -.07 | -2.03 | .04 | .71 | -.08 |
| No formal | | -.04 | -1.14 | .25 | 3.01 | -.04 |
| Unknown | | -.04 | -1.37 | .17 | 3.00 | -.05 |
