## Supplemental File 12_Table 3 for "Determinants of Exerciser Environmentalism to Lower Meat Consumption in the Practice of Climate Stewardship"

**Table 3**
Results of block five multiple regression analyses of the VBNTPB causal pathway two to reduce meat consumption for climate stewardship. (Two-Way Interaction).

| | b | β | *t* | *p* | S.E. | Partial correlation | 95% CI $R^2$ | Adj $R^2$ | df | Sig *F* change | Δ $R^2$ |
|---|---|---|---|---|---|---|---|---|---|---|---|
| *DV: Intention* | | | | | | | [.83-.87] | .85 | 3, 576 | <.001 | .197 |
| Group | .533 | .03 | 1.87 | .06 | .28 | .07 | | | | | |
| Gender | .391 | .02 | 1.39 | .16 | .28 | .05 | | | | | |
| Personal norm–Attitude | | .05 | 3.33 | .<.001 | .00 | .13 | | | | | |
| Personal norm | | .44 | 12.14 | <.001 | .04 | .45 | | | | | |
| Attitude | | .34 | 9.33 | <.001 | .04 | .36 | | | | | |
| Ascription of responsibility | | .08 | 2.97 | .00 | .04 | .12 | | | | | |
| Awareness of consequences | | .11 | 3.60 | <.001 | .03 | .14 | | | | | |
| Proenvironmental worldview | | .00 | .21 | .83 | .03 | .00 | | | | | |
| Biospheric | | .03 | 1.31 | .18 | .06 | .05 | | | | | |
| Altruistic | | -.03 | -1.83 | .06 | .06 | -.07 | | | | | |
| Egoistic | | .00 | .46 | .64 | .04 | .01 | | | | | |
| 28-37 | | .00 | .07 | .94 | .44 | .00 | | | | | |
| 38-47 | | .00 | .12 | .90 | .45 | .00 | | | | | |
| 48-57 | | -.00 | -.30 | .76 | .45 | -.01 | | | | | |
| 58-100 | | .01 | .58 | .55 | .48 | .02 | | | | | |
| Asian | | .00 | .17 | .86 | .69 | .00 | | | | | |
| Black | | .02 | 1.36 | .17 | .51 | .05 | | | | | |
| Mixed | | -.01 | -1.20 | .23 | .62 | -.05 | | | | | |
| Other | | .00 | .09 | .92 | .69 | .00 | | | | | |
| Technical/Community College | | -.01 | -.90 | .36 | .37 | -.03 | | | | | |
| Secondary | | -.00 | -.09 | .92 | .33 | -.00 | | | | | |
| No formal | | -.04 | -2.63 | .00 | 1.37 | -.10 | | | | | |
| Unknown | | -.01 | -.73 | .46 | 1.37 | -.03 | | | | | |
