## Supplemental File 13_Table 4 and 5 for "Determinants of Exerciser Environmentalism to Lower Meat Consumption in the Practice of Climate Stewardship"

**Table 4**
Results of block five multiple regression analyses of the VBNTPB causal pathway two to reduce meat consumption for climate stewardship. (Two-Way Interaction).

| | b | β | *t* | *p* | S.E. | Partial correlation | 95% CI $R^2$ | Adj $R^2$ | df | Sig *F* change | Δ $R^2$ |
|---|---|---|---|---|---|---|---|---|---|---|---|
| *DV: Intention* | | | | | | | [.84-.88] | .86 | 6, 573 | <.001 | .207 |
| Group | .602 | .03 | 2.16 | .29 | .27 | .09 | | | | | |
| Gender | .247 | .01 | .90 | .36 | .27 | .03 | | | | | |
| Personal norm–Attitude | | -.05 | -1.70 | .08 | .00 | -.07 | | | | | |
| Personal norm–Health concern | | -.00 | -.18 | .85 | .00 | -.00 | | | | | |
| Attitude–Health concern | | .14 | .14 | <.001 | .00 | .15 | | | | | |
| Personal norm | | .39 | 10.69 | <.001 | .41 | .40 | | | | | |
| Health concern | | .16 | 5.21 | <.001 | .51 | .21 | | | | | |
| Attitude | | .33 | 9.31 | <.001 | .04 | .36 | | | | | |
| Ascription of responsibility | | .07 | 2.48 | .01 | .04 | .10 | | | | | |
| Awareness of consequences | | .06 | 1.84 | .06 | .04 | .07 | | | | | |
| Proenvironmental worldview | | .00 | .21 | .82 | .03 | .00 | | | | | |
| Biospheric | | .02 | .89 | .37 | .06 | .03 | | | | | |
| Altruistic | | -.03 | -1.65 | .09 | .06 | -.06 | | | | | |
| Egoistic | | .01 | .75 | .45 | .03 | .03 | | | | | |
| 28-37 | | .00 | .47 | .63 | .43 | .02 | | | | | |
| 38-47 | | .01 | .58 | .55 | .44 | .02 | | | | | |
| 48-57 | | -.00 | -.12 | .90 | .44 | -.00 | | | | | |
| 58-100 | | .02 | .90 | .36 | .46 | .03 | | | | | |
| Asian | | -.01 | .50 | .61 | .67 | .02 | | | | | |
| Black | | .02 | .02 | .14 | .49 | .06 | | | | | |
| Mixed | | -.01 | -.91 | .36 | .61 | -.03 | | | | | |
| Other | | .00 | .07 | .93 | .66 | .00 | | | | | |
| Technical/Community College | | -.01 | -.84 | .40 | .36 | -.03 | | | | | |
| Secondary | | .01 | .58 | .56 | .32 | .02 | | | | | |
| No formal | | -.02 | -1.79 | .07 | 1.34 | -.07 | | | | | |
| Unknown | | -.01 | -.69 | .49 | 1.33 | -.00 | | | | | |

**Table 5**
Results of block five multiple regression analyses of the VBNTPB causal pathway two to reduce meat consumption for climate stewardship. (Three-Way Interaction).

| | b | β | *t* | *p* | S.E. | Partial correlation | 95% CI $R^2$ | Adj $R^2$ | df | Sig *F* change | Δ $R^2$ |
|---|---|---|---|---|---|---|---|---|---|---|---|
| *DV: Intention* | | | | | | | [.86-.89] | .87 | 7, 572 | <.001 | .215 |
| Group | .538 | .03 | 2.01 | .04 | .26 | .08 | | | | | |
| Gender | .261 | .01 | .89 | .32 | .26 | .04 | | | | | |
| Personal norm–Attitude-Health concern | | -.19 | -6.28 | <.001 | .00 | -.25 | | | | | |
| Personal norm–Attitude | | -.02 | -.90 | .36 | .00 | -.03 | | | | | |
| Personal norm–Health concern | | .00 | -.01 | .99 | .00 | .00 | | | | | |
| Attitude–Health concern | | .06 | 1.59 | .11 | .00 | .06 | | | | | |
| Personal norm | | .41 | 11.61 | <.001 | .04 | .43 | | | | | |
| Health concern | | .22 | 7.01 | <.001 | .05 | .28 | | | | | |
| Attitude | | .37 | 10.55 | <.001 | .04 | .40 | | | | | |
| Ascription of responsibility | | .08 | 3.03 | .00 | .03 | .12 | | | | | |
| Awareness of consequences | | .07 | 2.26 | .02 | .03 | .09 | | | | | |
| Proenvironmental worldview | | .01 | .54 | .58 | .03 | .02 | | | | | |
| Biospheric | | .02 | 1.01 | .31 | .06 | .04 | | | | | |
| Altruistic | | -.03 | -1.74 | .08 | .05 | -.07 | | | | | |
| Egoistic | | .01 | 1.19 | .23 | .03 | .05 | | | | | |
| 28-37 | | .01 | .98 | .32 | .42 | .04 | | | | | |
| 38-47 | | .02 | 1.25 | .21 | .42 | .05 | | | | | |
| 48-57 | | .00 | .12 | .90 | .43 | .00 | | | | | |
| 58-100 | | .03 | 1.60 | .10 | .45 | .06 | | | | | |
| Asian | | .01 | .78 | .43 | .65 | .03 | | | | | |
| Black | | .02 | 1.56 | .11 | .47 | .06 | | | | | |
| Mixed | | -.01 | -.66 | .50 | .59 | -.02 | | | | | |
| Other | | .00 | .22 | .82 | .64 | .00 | | | | | |
| Technical/Community College | | -.00 | -.50 | .61 | .35 | -.02 | | | | | |
| Secondary | | -.03 | .77 | .43 | .31 | .03 | | | | | |
| No formal | | -.00 | -2.20 | .02 | 1.30 | -.09 | | | | | |
| Unknown | | -.00 | -.38 | .69 | 1.29 | -.01 | | | | | |
