## Supplemental File 14_Table 6 and 7 for "Determinants of Exerciser Environmentalism to Lower Meat Consumption in the Practice of Climate Stewardship"

**Table 6**
Results of block five multiple regression analyses of the VBNTPB causal pathway three to reduce meat consumption for climate stewardship. (Two-Way Interaction).

| | b | β | *t* | *p* | S.E. | Partial correlation | 95% CI $R^2$ | Adj $R^2$ | df | Sig *F* change | Δ $R^2$ |
|---|---|---|---|---|---|---|---|---|---|---|---|
| *DV: Intention* | | | | | | | [.84-.88] | .86 | 6, 573 | <.001 | .204 |
| Group | .456 | .02 | 1.63 | .10 | .27 | .06 | | | | | |
| Gender | .498 | .02 | 1.81 | .07 | .27 | .07 | | | | | |
| Personal norm–Attitude | | .04 | 1.78 | .07 | .00 | .07 | | | | | |
| Personal norm–Social norm | | .02 | .55 | .57 | .00 | .02 | | | | | |
| Social norm–Attitude | | -.00 | -.04 | .96 | .01 | -.00 | | | | | |
| Personal norm | | .38 | 10.15 | <.001 | .04 | .39 | | | | | |
| Social norm | | .12 | 5.19 | <.001 | .04 | .21 | | | | | |
| Attitude | | .32 | 8.62 | <.001 | .04 | .33 | | | | | |
| Ascription of responsibility | | .07 | 2.62 | .00 | .04 | .10 | | | | | |
| Awareness of consequences | | .11 | 3.84 | <.001 | .03 | .15 | | | | | |
| Proenvironmental worldview | | .00 | .23 | .81 | .03 | .01 | | | | | |
| Biospheric | | .03 | 1.16 | .24 | .26 | .04 | | | | | |
| Altruistic | | -.03 | -1.45 | .14 | -.03 | -.06 | | | | | |
| Egoistic | | .00 | -.00 | .99 | .00 | .00 | | | | | |
| 28-37 | | .00 | .44 | .65 | .00 | .00 | | | | | |
| 38-47 | | .00 | .12 | .89 | .43 | .01 | | | | | |
| 48-57 | | .00 | .15 | .87 | .44 | .00 | | | | | |
| 58-100 | | .01 | .49 | .62 | .45 | .00 | | | | | |
| Asian | | .00 | .28 | .77 | .68 | .02 | | | | | |
| Black | | .02 | 1.42 | .15 | .50 | .01 | | | | | |
| Mixed | | -.01 | -1.05 | ..29 | .61 | .05 | | | | | |
| Other | | .00 | .52 | .60 | .67 | -.04 | | | | | |
| Technical/Community College | | -.01 | -.61 | .53 | .37 | -.02 | | | | | |
| Secondary | | .00 | .14 | .88 | .32 | .00 | | | | | |
| No formal | | -.03 | -1.98 | .04 | 1.36 | -.08 | | | | | |
| Unknown | | -.01 | -1.17 | .24 | 1.34 | -.04 | | | | | |

**Table 7**
Results of block five multiple regression analyses of the VBNTPB causal pathway three to reduce meat consumption for climate stewardship. (Three-Way Interaction).

| | b | β | *t* | *p* | S.E. | Partial correlation | 95% CI $R^2$ | Adj $R^2$ | df | Sig *F* change | Δ $R^2$ |
|---|---|---|---|---|---|---|---|---|---|---|---|
| *DV: Intention* | | | | | | | [.85-.88] | .87 | 7, 572 | <.001 | .213 |
| Group | .406 | .02 | 1.50 | .13 | .27 | .06 | | | | | |
| Gender | .507 | .02 | 1.90 | .05 | .26 | .07 | | | | | |
| Personal norm–Attitude-Social norm | | -.16 | -6.18 | <.001 | .00 | -.25 | | | | | |
| Personal norm–Attitude | | .02 | .99 | .32 | .00 | .04 | | | | | |
| Personal norm–Social norm | | .05 | 1.34 | .17 | .00 | .05 | | | | | |
| Attitude–Social norm | | -.02 | -5.2 | .60 | .01 | .06 | | | | | |
| Personal norm | | .38 | 10.42 | <.001 | .04 | -.02 | | | | | |
| Social norm | | .20 | 7.69 | <.001 | .05 | .30 | | | | | |
| Attitude | | .35 | 9.82 | <.001 | .04 | .38 | | | | | |
| Ascription of responsibility | | .08 | 3.02 | .00 | .03 | .12 | | | | | |
| Awareness of consequences | | .13 | 4.42 | <.001 | .03 | .18 | | | | | |
| Proenvironmental worldview | | .01 | .64 | .52 | .03 | .02 | | | | | |
| Biospheric | | .02 | 1.06 | .28 | .06 | .04 | | | | | |
| Altruistic | | -.02 | -1.28 | .19 | .05 | -.05 | | | | | |
| Egoistic | | .00 | .33 | .74 | .03 | .01 | | | | | |
| 28-37 | | .01 | .90 | .36 | .42 | .03 | | | | | |
| 38-47 | | .01 | .51 | .61 | .43 | .02 | | | | | |
| 48-57 | | .00 | .29 | .76 | .43 | .01 | | | | | |
| 58-100 | | .02 | 1.04 | .29 | .46 | .04 | | | | | |
| Asian | | .00 | .38 | .70 | .66 | .01 | | | | | |
| Black | | .02 | 1.36 | .06 | .48 | .07 | | | | | |
| Mixed | | -.01 | -.90 | .36 | .59 | -.03 | | | | | |
| Other | | .01 | .72 | .47 | .65 | .03 | | | | | |
| Technical/Community College | | -.01 | -.79 | .42 | .35 | -.03 | | | | | |
| Secondary | | -.00 | -.22 | .81 | .31 | -.01 | | | | | |
| No formal | | -.02 | -1.86 | .06 | 1.31 | -.07 | | | | | |
| Unknown | | -.01 | -.84 | .39 | 1.30 | -.03 | | | | | |
