## Supplemental File 15_Table 8 for "Determinants of Exerciser Environmentalism to Lower Meat Consumption in the Practice of Climate Stewardship"

Results of multiple regression indirect effects analyses of the VBN and VBNTPB (1-3) causal pathways to reduce meat consumption for climate stewardship.

| | Independent variable | Mediator | Dependent variable | Indirect effect | SE | 95% confidence interval for indirect effect (Bias corrected and accelerated). | |
|---|---|---|---|---|---|---|---|
| | | | | | | Lower | Upper |
| VBN casual pathway | | | | | | | |
| | BV | PWV | AC | .18*** | .00 | .578 | .580 |
| | AV | PWV | AC | .07*** | .00 | .578 | .580 |
| | EV | PWV | AC | .03*** | .00 | .578 | .580 |
| | PWV | AC | AR | .03*** | .00 | .100 | .103 |
| | AC | AR | PN | .03*** | .00 | .639 | .642 |
| | AR | PN | IN | .31*** | .00 | .780 | .781 |
| VBNTPB casual pathway 1 (two-way interaction) | | | | | | | |
| | AR | PN | IN | .20*** | .00 | .501 | .503 |
| | AR | AT | IN | .15*** | .00 | .445 | .447 |
| | AR | AT-PN | IN | .02*** | .00 | .009 | .009 |
| VBNTPB casual pathway 2 (two-way interaction) | | | | | | | |
| | AR | PN | IN | .17*** | .00 | .443 | .445 |
| | AR | AT | IN | .15*** | .00 | .430 | .432 |
| | AR | HC | IN | .07*** | .00 | .262 | .265 |
| | AR | AT-PN | IN | -.02*** | .00 | -.008 | -.008 |
| | AR | HC-AT | IN | .06*** | .00 | .028 | .028 |
| | AR | HC-PN | IN | -.00*** | .00 | -.001 | -.001 |
| VBNTPB casual pathway 2 (three-way interaction) | | | | | | | |
| | AR | PN | IN | .18*** | .00 | .468 | .470 |
| | AR | AT | IN | .16*** | .00 | .478 | .481 |
| | AR | HC | IN | .10*** | .00 | .358 | .360 |
| | AR | AT-PN | IN | -.01*** | .00 | -.004 | -.004 |
| | AR | PN | IN | .18*** | .00 | .468 | .470 |
| | AR | AT | IN | .16*** | .00 | .478 | .481 |

| | | | | | | | |
|---|---|---|---|---|---|---|---|
| | AR | HC | IN | .10*** | .00 | .358 | .360 |
| VBNTPB casual pathway 3 (two-way interaction) | | | | | | | |
| | AR | PN | IN | .19*** | .00 | .431 | .433 |
| | AR | AT | IN | .18*** | .00 | .407 | .409 |
| | AR | SN | IN | .10*** | .00 | .231 | .234 |
| | AR | SN/PN | IN | .00*** | .00 | .004 | .005 |
| | AR | SN/AT | IN | -.00*** | .00 | -.001 | -.001 |
| | AR | AT/PN | IN | .01*** | .00 | .007 | .007 |
| VBNTPB casual pathway 3 (three-way interaction) | | | | | | | |
| | AR | PN | IN | .17*** | .00 | .428 | .431 |
| | AR | AT | IN | .16*** | .00 | .456 | .458 |
| | AR | SN | IN | .09*** | .00 | .381 | .383 |
| | AR | SN/PN | IN | .02*** | .00 | .011 | .011 |
| | AR | AT/PN | IN | .01*** | .00 | .004 | .004 |
| | AR | SN/AT | IN | -.00*** | .00 | -.005 | -.005 |
| | AR | SN/AT/PN | IN | -.07*** | .00 | -.003 | -.003 |

*Note:* BV = biospheric value orientation, AV = altruistic value orientation, EV = egostic value orientation, PWV = proenvironmental worldview, AC = awareness of consequences, AR = ascription of responsibility, PV = personal norm, AT = attitude, HC = health concern, SN = social norm.
***$p$ < .001
